# Minimizing biological risk for novel inhibitory drug targets: One knockout is all you need

**DOI:** 10.1101/2024.06.19.24309116

**Authors:** Alan Dimitriev, Lynne-Marie Postovit, Amber L. Simpson, Gane Ka-Shu Wong

## Abstract

We argue that biological risk for novel inhibitory drug targets can be minimized, almost eliminated, by a computational analysis of the healthcare records and DNA sequences in resources like UK Biobank or All-of-Us. The key insight is that an inhibitory drug is functionally equivalent to a loss-of-function (LOF) variant in the targeted gene. It is a special case of what has been called an “experiment of nature”. To demonstrate, we considered all available clinical trials (58 in total) and inhibitory drugs (15 in total) for 5 cardiovascular drug targets: PCSK9, APOC3, ANGPTL3, LPA, and ASGR1. The results were shocking. Every biomarker assessed in these clinical trials was successfully predicted, i.e. directionality and proportionality of effect, but not the magnitude since that varies with dosage. This concept has not been widely adopted because geneticists believe that homozygous LOFs, which are exceedingly rare, would be needed to observe a significant phenotypic effect from most genetic knockouts. Our study shows that, to the contrary, given a sufficiently large biobank, counting both carriers and non-carriers, heterozygous LOFs alone can inform drug development.

## Introduction

The development of novel drug therapies is a resource-intensive process where a majority of the expenses are due to the amortized costs of attrition, with the total research and development cost per new drug in 2016 being estimated to be $2.5 billion^1,2^. The primary contributor to this financial crisis is the high rate of failure, nominally 90%, and predominantly attributed to a lack of efficacy as opposed to safety^3–7^. Preclinical validation of drug targets has historically relied on animal models and/or cell lines. These approaches face inherent limitations when translating results to humans^8,9^. Retrospective analyses of drug development processes has highlighted that drug targets with human genetics evidence are more than twice as likely to reach approval as those without^10–15^, a reminder that the best model of human biology is a human subject, not an animal model or cell line. Here, we will demonstrate how DNA sequencing, combined with electronic health records, can de-risk the drug development process by reliably predicting clinical trial outcomes.

Conceptually, our method is what Plenge *et al*. called an “experiment of nature” in which a naturally occurring human condition ***directly*** mimics the effect of the drug, as opposed to the traditional discovery process that focuses on understanding the mechanistic causes of disease^10^. We will consider the special case of an inhibitory drug, where the mode-of-action is to disrupt the function of a targeted gene/protein. Drugs of this nature include antisense oligonucleotides (ASO), monoclonal antibodies, small interfering RNAs (siRNAs), traditional small molecules, and in some instances small peptides. The corresponding human condition would be any number of naturally occurring loss-of-function (LOF) variants in the targeted gene. These LOFs are functionally equivalent to the inhibitory drug, insofar as they exert a qualitatively similar effect on biomarker changes associated with intermediate phenotypes and on disease progression. As such, they have been called “human knockouts as models of drug action” (HKMD)^16^. Quantitative differences between genetic and clinical studies are expected to stem from the zygosity of the LOF variants and from the dosage of the drug in question.

Although ideal, preclinical models of this nature are only practical when we can identify sufficiently many human subjects with the requisite variants. This is primarily feasible for heterozygous LOFs (one knockout), because homozygous LOFs (two knockouts) are extremely rare^17^, at least in outbred populations. Such methods would be of limited utility if homozygous LOFs were required, as it has been shown that even if we were to sequence everybody worldwide, we would never get homozygous LOFs for many genes^18–20^. Conversely, relying on the plentiful heterozygous LOFs that can be found in currently available biobanks is problematic, because most genes are haplosufficient (Mendelian recessive) and there is little to no organismal phenotypic difference between the homozygous wild type and a heterozygous LOF^21–24^. This conception of recessiveness dates back to Mendel, and the mystery of how it works inspired two of the giants of early 20^th^ century genetics to suggest compensatory mechanisms. Fisher’s 1928 proposal was based on evolutionary selection for genetic modifiers^25^. Wright’s 1934 counterproposal was predicated on the redundancy of cellular functions^26^. Regardless of the explanation, it is the prevailing orthodoxy that heterozygous LOFs tend not to be informative, and this underlies the interest in specialized populations (e.g. consanguineous, bottlenecked) where homozygous LOFs are more common^27,28^.

Despite the above concern, we set out to ascertain the feasibility of using currently available human genetic/medical data to perform preclinical validation of novel drug targets. Specifically, we utilized the UK Biobank to identify carriers of genetic variants that are predicted to cause LOF in five genes (*PCSK9, APOC3, ANGPTL3, LPA, ASGR1*) that have been reported to improve a cardiovascular biomarker and to be protective against ischemic heart disease. All have been targeted by drugs in clinical trials that are at various stages of completion. We contrasted the changes in biomarker distribution between predicted loss-of-function (pLOF) variant carriers and the very large number of non-carriers in the UK Biobank. Then we compared our results to the differences in biomarker distribution observed in clinical trials between individuals undergoing inhibitory drug therapy versus those receiving a placebo. From our genetic study, we consistently observed the same directionality and proportionality of effect as was reported in the clinical trials, demonstrating the ability of human genetics to be used for preclinical validation.

## Methods

The UK Biobank has approval from the North West Multi-Centre Research Ethics Committee as a Research Tissue Bank (RTB). This approval means that researchers do not require separate ethical clearance by their institution and can operate under the RTB approval. Our research was conducted under Application Number 85442. The UK Biobank is a prospective cohort study that over a four-year period successfully recruited over 500,000 volunteer participants in an effort to investigate the risk factors for major diseases of middle and old age. The UK Biobank was uniquely positioned for this research project as it offers access to both genomic data and blood biomarker measurements for a large number of participants. Following previous work utilizing the UK Biobank, we performed quality control on biobank participants by excluding individuals marked as having sex chromosome aneuploidy (Data-Field 22019, UKB - 651 participants), a mismatch between self-reported and genetically determined sex (Data-Field 31 – UKB, Data-Field 22001 – UKB, 372 subjects), outliers for heterozygosity or missing rate (Data-Field 22027, UKB - 968 participants), and those who were not used in the genetic principal component analysis performed by the UK Biobank to determine genetic ancestry (Data-Field 22020, UKB - 95,361 participants)^29^.

We stratified patients by the presence of the flag ‘Caucasian’ in Data-Field 22006. This was done to reduce genetic variation noise and minimize the effects of population stratification, as a majority of the UK Biobank participants are Caucasian. This flag indicates samples who self-identified as ‘White British’ according to Data-Field 21000 and have similar genetic ancestry based on a principal components analysis of the genotypes. This resulted in a study population of 337,083 participants.

We defined cases as individuals having the International Statistical Classification of Diseases and Related Health Problems, 10th edition (ICD-10), diagnostic codes for Ischemic Heart Diseases (I20-I25), which include: angina pectoris (I20), acute myocardial infarction (I21), subsequent myocardial infarction (I22), certain current complications following acute myocardial infarction (I23), other acute ischaemic heart diseases (IHD) (I24), and chronic ischemic heart disease (I25). Case participants were identified at the time of this analysis (July 2022) as those who had any of the above ICD-10 codes listed as a primary, secondary, or tertiary diagnoses or as a cause-of-death diagnosis.

The UK Biobank provides blood biochemistry measures on 30 different blood biomarkers (a full list of biomarkers can be found at https://biobank.ndph.ox.ac.uk/showcase/label.cgi?id=17518). We performed our analysis on all 30 biomarkers for each gene and report on the significant associations observed.

When working with genetic variants the UK Biobank recommends that a single variant-level filter be applied requiring that at least 90% of all genotypes for a given variant — independent of variant allele zygosity — have a read depth of at least 10 (i.e., DP ≥ 10). When this filter is applied to the UKB whole exome sequence (WES) 200k data prior to association analysis, the results are largely devoid of the spurious hits. Variants from pVCF files were filtered through BCFtools following the procedure outlined by Ghouse et al., where in we filtered out variants with a genotype quality of less than 20, a genotype depth of less than 10, or that had missing genotype percentage over 10%^30^. Variants were functionally annotated using SnpEff version 5.0e (build 2021-03-09 06:01)^31^.

The high confidence (HC) set of pLOF variants includes only variants annotated to be LOF according to SnpEff, based on the following criteria set out by MacArthur et al.: stop codon-introducing (nonsense), splice site-disrupting single-nucleotide variants (SNVs), insertion/deletion (indel) variants predicted to disrupt a transcript’s reading frame, or larger deletions removing either the first exon or more than 50% of the protein-coding sequence of the affected transcript^17,32^. Of note, the variants that meet these criteria but are within the first and last 5% of the gene region were excluded. Variants captured by our analysis can be seen in Supplementary Tables 3-7.

Combined Multivariate and Collapsing (CMC) Burden association testing was done using the standalone RVTESTS package, a popular and flexible package due to its support for large-scale biobank data^33,34^. Burden tests were performed using gene regions as the grouping parameter under the assumption that all pLOF variants have the same directionality of effect. All models were adjusted for age at enrollment, sex, and genetic ancestry (as quantified by the first five principal components), and non-normally distributed variables were log-transformed (these being: alanine aminotransferase, c-reactive protein, direct bilirubin, gamma glutamyltransferase, lipoprotein A, oestradiol, rheumatoid factor, total bilirubin, and triglycerides).

To compare our genetic results with the relevant clinical trials, we searched www.clinicaltrials.gov. The full list of what we found can be seen in Supplementary Tables 8-12. We believe this to be an exhaustive collection of all publicly available clinical trials for all inhibitory drugs created for our five targets. It includes 58 trials for 15 drug compounds broken down by target as 30 trials for 5 drugs consisting of 3 modalities (monoclonal antibody, siRNA, and macrocyclic peptide) targeting PCSK9, 10 trials for 3 drugs consisting of 2 modalities (ASO and siRNA) targeting APOC3, 12 trials for 3 drugs consisting of 3 modalities (ASO, monoclonal antibody, and siRNA) targeting ANGPTL3, 5 trials for 3 drugs consisting of 2 modalities (ASO and siRNA) targeting LPA, and the single available trial of a monoclonal antibody targeting ASGR1.

## Results

We considered all 30 blood biomarkers available in the UK Biobank and reported on those observed to have statistically significant changes in distribution between carriers and non-carriers of pLOF alleles. For all observed distributions, see Supplementary Table 1. We also compared our results to those reported by previous genetic studies, as can be seen in Supplementary Table 2. Comparisons between our genetic studies and the published clinical studies for each gene can be seen in Table 1.

**Table 1.**
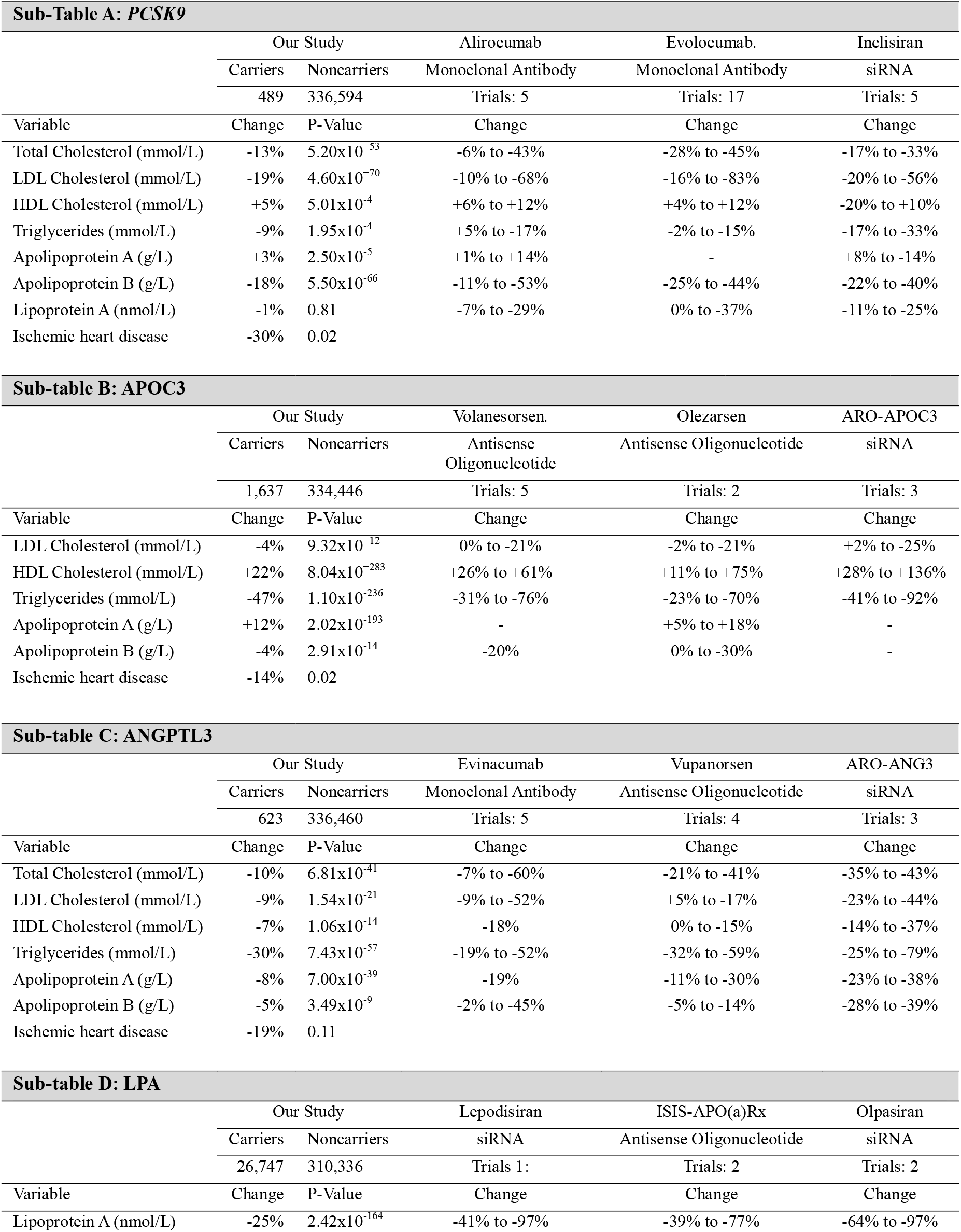
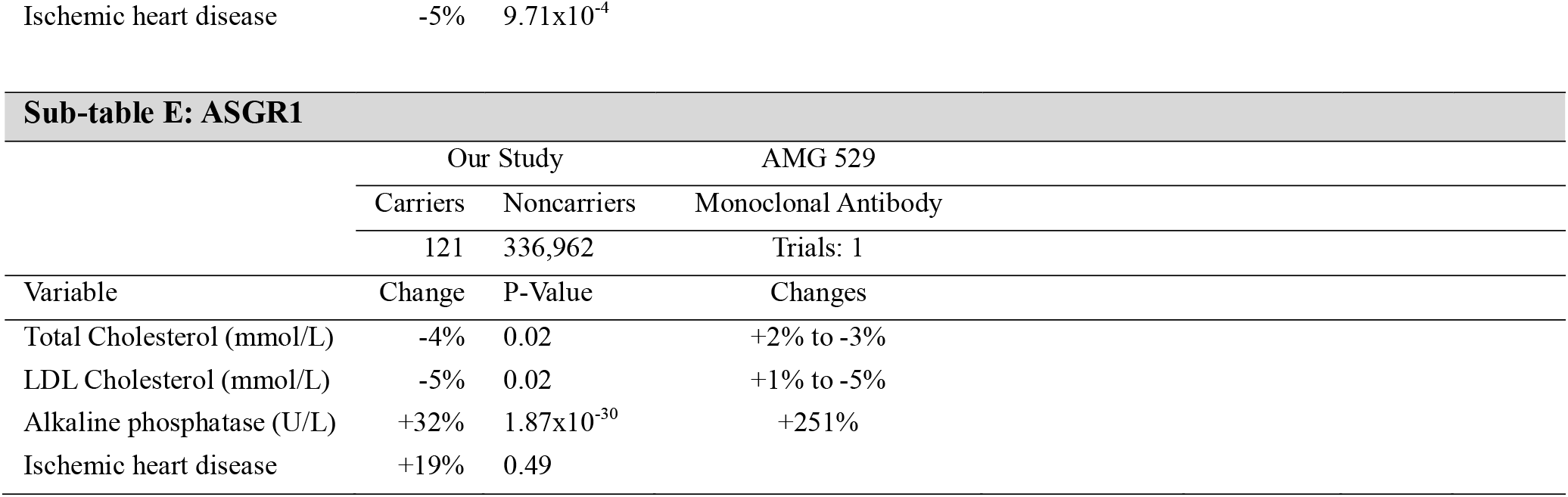
Comparison between predicted loss-of-function variants and clinically reported effects of inhibitory drugs on the distribution of blood biomarkers for Ischemic Heart Disease.

### PCSK9

We identified 489 carriers of 41 unique high-confidence *PCSK9* pLOF variants, equating to a carrier frequency of 0.15%. Within this cohort, we observed one individual who was multi-allelic for pLOF variants, and no homozygous carriers at all. We observed that carriers of a LOF variant in the *PCSK9* gene had significant reductions compared to noncarriers in plasma levels of low-density lipoprotein cholesterol (LDL-C) (−19%, P=4.60×10^−70^), apolipoprotein B (apoB) (−18%, P=5.50×10^−66^), total cholesterol (−13%, P=5.20×10^−53^), and triglycerides (−9%, P=1.95×10^−4^), while having increases in plasma levels of high-density lipoprotein cholesterol (HDL-C) (+5%, P=5.01×10^−4^), and apolipoprotein A (apoA) (+3%, 2.50×10^−4^). Lastly, we observed a protective effect against IHD from the presence of a *PCSK9* LOF allele, although the statistical significance is weak.

We compared our findings on the effect of *PCSK9* pLOF variants to thirty prior clinical studies investigating therapeutic inhibitors targeting *PCSK9*. These studies include notable trials such as the 2017 trial on evolocumab by Sabatine *et al*.^35^, the 2018 trial on alirocumab by Schwartz *et al*.^36^, and the 2020 trial on inclisiran by Ray *et al*.^37^ The strongest signal observed in our study was the reduction in plasma LDL-C, consistently aligning with outcomes from clinical trials where therapeutic inhibition of *PCSK9* demonstrated a decrease in mean plasma LDL-C levels across all trials. The proportionality of this result is also consistent as LDL-C saw the highest reported percent change of any biomarker in each clinical trial, with maximum observed reductions ranging from -56% to -83%. While the quantitative strength of our observed reduction is less than that of most trials, this is expected as our study focused on heterozygous carriers of *PCSK9* pLOF variants.

A promising aspect of our study is how our results align with changes in the distribution of all clinical variables. In all trials where they were reported, both total cholesterol and apolipoprotein B levels saw percent change decreases ranging from -6% to -45% and -11% to -53%, respectively, which is mirrored in our results as we observed reductions of -13% in mean total cholesterol levels and -18% in mean apolipoprotein B levels, both at high significance thresholds.

Comparison of our results for HDL-C, triglycerides, and apolipoprotein A with previous clinical trials adds further confidence to our proof of concept. The prevailing trend in clinical trial results was that therapeutic inhibition of PCSK9 resulted in reductions in plasma triglyceride levels, and slight increases in HDL-C and apolipoprotein A levels. However, for each biomarker there was one trial that reported the alternative effect. Our analysis of pLOF variants resulted in reported directionality of biomarker changes in line with the majority of clinical trials for each of these biomarkers, showcasing a potential advantage that large scale genetic analysis can offer over the variability of clinical trials with a smaller sample population. The only deviation from this trend is that it has been reported in previous clinical trials that mean plasma lipoprotein A levels decreased in study participants who received therapeutic inhibition. We did not observe any significant change in the plasma levels of lipoprotein A between carriers and non-carriers of pLOF variants.

Our results highlight that, despite the quantitative strength of the change in biomarker distribution being generally lower than those reported in clinical trials, heterozygous carriers of pLOF variants can be used to observe the same directionality of effect as gene-specific inhibition in a clinical trial.

### APOC3

We identified 1,637 carriers of seven unique high-confidence *APOC3* pLOF variants (equating to a carrier frequency of 0.49%), all of whom were heterozygous carriers. Our analyses revealed that carriers of a LOF variant in the *APOC3* gene had significant increases in plasma levels of HDL-C (+22%, P=8.04×10^−283^) and apolipoprotein A (+12%, P= 2.02×10^−193^), as well as significant reductions in plasma levels of LDL-cholesterol (−4%, P=9.32×10^−12^), triglycerides (−47%, P=1.10×10^−236^), and apolipoprotein B (−4%, P=2.91×10^−14^). Once again, we observed a protective effect against IHD from the presence of a *APOC3* LOF allele (−14%, P=0.02), but the statistical significance is weak.

The largest change in biomarker distribution due to the presence of an *APOC3* pLOF allele was in mean levels of plasma triglycerides, which is consistent with previous clinical investigations into *APOC3* inhibitors by Gouni-Berthold *et al*., Schwabe *et al*., and Alexander *et al*., on volanesorsen, ARO-APOC3, and olezarsen, respectively^38–40^. These trials reported reductions of mean plasma triglyceride levels by - 53% to -70%. We expanded our comparison to a total of ten clinical trials and noted that in all cases inhibition of *APOC3* resulted in a reduction of plasma triglycerides ranging from -23% to -92%. Across all ten trials, where reported, therapeutic inhibition of *APOC3* resulted in observed increases of plasma HDL-C levels ranging from +11% to +136%, which is mirrored in our genetic study, where we observed a significant increase in plasma HDL-C levels. Barring one study that reported a +2% increase in mean LDL-C levels after therapy, our observed reductions in LDL-C and apolipoprotein B levels are consistent with all major clinical trials for *APOC3* inhibitors, and our observed increase in apolipoprotein A levels is consistent with clinical trials where reported.

When compared to the results of previous clinical trials, our genetic results from the UK Biobank align across every biomarker in terms of predicting not only the directionality of the change in blood biomarker distribution caused by inhibition, but also the proportionality of the relative effect on each specific biomarker. As with our results for *PCSK9*, we were able to clearly demonstrate the effect of gene specific inhibition without having to do a clinical trial.

### ANGPTL3

We identified 623 carriers of 33 unique high-confidence *ANGPTL3* pLOF variants, equating to a carrier frequency of 0.18%. We found one individual homozygous for a pLOF variant, with the rest being heterozygous carriers. Our analyses revealed that carriers of a LOF variant in the *ANGPTL3* gene had significant reductions in plasma levels of total cholesterol (−10%, P=6.81×10^−41^), LDL-C (−9%, P=1.54×10^−21^), HDL-C (−7%, P=1.06×10^−14^), triglycerides (−30%, P=< 7.43×10^−57^), apolipoprotein A (−8%, P=< 7.00×10^−39^), and apolipoprotein B (−5%, P=3.49×10^−9^). Furthermore, we observed a protective effect against IHD from the presence of an *ANGPTL3* LOF allele, although this effect was not statistically significant.

We compared our results to those reported by twelve prior clinical trials on the monoclonal antibody evinacumab, the ASO vupanorsen, and the siRNA ARO-ANG3. This includes, but is not limited to, a 2020 trial on vupanorsen by Bergmark et al.^41^, and a 2020 trial on evinacumab by Harada-Shiba *et al*.^42^ Across all trials, plasma levels of triglycerides and total cholesterol saw the largest reductions, which is consistent with our observed results. Every clinical trial for *ANGPTL3* inhibitors reported reductions in plasma levels of LDL-C, HDL-C, apoA, and apoB as a result of therapeutic intervention. This trend is mirrored in our genetic results. Once again, we were able to successfully observe the directionality of the change in biomarker distribution that arises due to therapeutically induced LOF.

### LPA

We identified 26,747 carriers of 111 unique high-confidence *LPA* pLOF, equating to a carrier frequency of 7.93%. Unique among our 5 targeted gene, we observed 485 carriers who were homozygous for a pLOF variant. Our analyses revealed that carriers of a LOF variant in the *LPA* gene had a significant reduction in plasma levels of lipoprotein A (−25%, P=2.42×10^−164^); however, we did not observe a significant or high impact change for any other blood biomarker that we analyzed. We did observe a protective effect against IHD from the presence of an *LPA* LOF allele (−5%, P=9.71×10^−4^).

A recent clinical trial on the siRNA lepodisiran targeting *LPA* by Nissen *et al*. reported reductions of plasma levels of lipoprotein A by -41% to -97%^43^. Clinical trials on the antisense oligonucleotide ISIS-APO(a)Rx and the siRNA olpasiran have reported similar ranges of plasma lipoprotein A reductions, ranging from -39% to -97%^44,45^. The key takeaway from the two latter studies was that both Tsimikas *et al*.^44^ and Koren *et al*.^45^ reported that none of the other lipid concentrations that we assessed, aside from lipoprotein A, saw significant change in their distribution when compared to the placebo group. This is entirely consistent with our results.

We investigated the distributions of homozygous carriers independently but found that only 6 individuals who were homozygous for a pLOF variant had associated plasma Lp(a) measurement. This is in contrast to 15,324 of the 26,747 heterozygous carriers and 240,203 of the 310,336 noncarriers who had an associated Lp(a) measurement. As biomarkers go, Lp(a) is an outlier with a history of being inconsistently measured, and this has other implications that will be raised in the discussions.

### ASGR1

We identified 121 carriers of 21 unique high-confidence *ASGR1* pLOF variants, equating to a carrier frequency of 0.04%, all of which were heterozygous carriers. Our analyses revealed that carriers of a LOF variant in the *ASGR1* gene had reductions in plasma levels of total cholesterol (−4%, P=0.02), LDL-C (−5%, P=0.02) and apolipoprotein B (−4%, P=0.01), as well as a significant increase in plasma alkaline phosphatase (+32%, P=1.87×10^−30^). It should be noted how weak the p-value significances of the first two changes are, when compared to the other biomarkers and genes in our study. Lastly, we did not observe a statistically significant protective effect against IHD from the presence of an *ASGR1* LOF allele; if anything, it made things worse.

While the statistical interpretation of our results for *ASGR1* is limited by a smaller sample size, it does indicate that inhibition of *ASGR1* might not have as consistent of a cholesterol lowering effect as hypothesised. Interestingly, while we did not observe a strong relationship regarding the cholesterol reducing effect of *ASGR1* inhibition, we did observe a 32% (P=1.87×10^−30^) increase in plasma levels of alkaline phosphatase. A phase 1 study of monoclonal antibody AMG 529 by Janiszewski *et al*. reported that, while the therapy was tolerated, it resulted in dose related increases in alkaline phosphatase (+251%) but, notably, no dose related effect on lipid or apolipoprotein measurements^46^. *ASGR1* stands apart from the previous target genes we investigated as it has limited clinical data to compare against; even so, the same directionality of change on biomarker distribution due to clinical inhibition can be observed in the prospective cohort population using pLOF variants, specifically the marked increase in liver enzymes but limited change in lipid levels.

## Discussion

To demonstrate how to de-risk the drug development process for inhibitory targets, we analyzed the phenotypic effects that pLOF in five target genes had on distributions of cardiovascular blood biomarkers. It should be noted that, for many common diseases, and on the time-scale of a clinical trial that is conducted for initial regulatory approval, biomarkers are the only practical measurables. There is never enough time to assess a long-term disease outcome. Our study was based on a Caucasian population of 337,083 UK Biobank participants. We identified pLOF variant carriers for each target gene and performed multivariate and collapsing burden tests to evaluate the association between LOF and phenotypic expression. Our study validated previously reported genetic associations between LOF carrier status and cardiovascular biomarker levels. Significant associations included reduced levels of LDL-C (*PCSK9, APOC3, ANGPTL3*), reduced levels of plasma triglycerides (*PCSK9, APOC3, ANGPTL3*), and reduced levels of lipoprotein A (*LPA*). We compared our genetic results to relevant clinical trials of inhibitory drugs and demonstrated that, for each target gene, we were able to successfully mirror the directionality and proportionality of change in blood biomarker distributions caused by therapeutic inhibition. Our analyses predicted not just one biomarker but the whole spectrum of biomarkers associated with the various intermediate phenotypes. The consistency of this approach is remarkable, given that between the clinical trials and our genetic studies, and across all target genes, there was not one significant deviation of the reported directionality of change for any biomarker that was considered. No false positives and no false negatives, to the extent that the same biomarker was assessed in both the clinical trial and our genetic study. This result is strengthened by the fact that our conclusions hold irrespective of the inhibition mechanism used in the clinical trial, whether that mechanism is ASO, monoclonal antibodies, siRNA, or small peptides. The only perceived exception to this assertion was that some clinical trials of PCSK9 inhibitors reported therapy induced reductions in mean levels of lipoprotein A, whereas we observed no significant change. However, it should be noted that Lp(a) levels are highly variable in terms of size and concentration, with a more than 1000-fold variation between individuals^47^. There exists the potential that, unless the observed change is very large, it will be challenging to detect a signal in the distribution of Lp(a) between carriers and non-carriers of pLOF variants.

The agreement reported here is a testament to how well these particular drugs inhibit the target gene/protein, without off-target binding, and the fact they are delivered to the target site at the appropriate concentrations. In other words, these molecules satisfy the binding and delivery requirements of any good drug. LOF-based validation addresses a different issue. Is the target itself a good choice? Our method addresses the biological risk of a novel inhibitory target, not the technological risk of the drug molecule itself. Because the assessment uses human data (not animal models or cell lines) that ***directly*** implicates a particular target with the desired phenotype, there is every reason to believe that it will be reliable. So, while LOF-based validation is not by itself sufficient to ensure clinical trial success, it should perhaps be a necessary condition for a decision to proceed onto the earliest stages of clinical development. This would help avoid wasted efforts on a molecule that is destined to fail. Lastly, since our analysis on the UK Biobank was restricted to Caucasians, the predicted outcomes may not be relevant for another population. Such concerns can be allayed by repeating the analysis on more genetically diverse resources like the All-of-Us^48^ databank.

To better understand why this approach works, consider the remarkably small p-values for the significance of the differences in the distribution of blood biomarkers between carriers and non-carriers of pLOF variants. This is especially notable when compared to the clinical trials. It is not an artifact of our method. A previous investigation by Ghouse *et al*. into the impact of *PCSK9* LOF variants on the distribution of glycemic biomarkers reported signals of similar strength using a subset of our UK Biobank data^30^. It has previously been highlighted that, with a sufficiently large sample size (and notice how the non-carriers in our study number in the many hundreds of thousands), a significant p-value is likely to be found even when the difference between groups is small^49^. The result to focus on is not just the statistical significance of the change observed, but also the magnitude of that change. For example, the primary biomarkers targeted by inhibitory drugs for *PCSK9* and *APOC3* are LDL-C and triglycerides, respectively. Our genetic studies found reductions of (−19%, P=4.60×10^−70^) and (−47%, P=1.10×10^−236^), respectively. With the magnitude and statistical significance of these genetic results, the likelihood that similar results would be observed in a clinical trial is very high. Conversely, for our weaker genetic findings, inconsistent results were occasionally seen among the clinical trials.

What is especially noteworthy is the fact that all of the targets we considered are haplosufficient (Mendelian recessive) genes, with estimated LOEUF (LOF observed / expected upper bound fraction, as reported in gnomAD) of 1.01, 0.97, 0.91, 1.05 and 0.76 for *PCSK9, APOC3, ANGPTL3, LPA*, and *ASGR1*, respectively^50^. Given how organismal phenotypic differences are expected to be small for heterozygous LOFs in recessive genes, this is probably why the magnitude of the biomarker changes in our genetic studies were generally smaller than what was reported in the clinical trials. Nonetheless, a reliable signal was detected in all instances. It is also important to distinguish between organismal phenotypes that are inevitably impacted by many other genes and environmental factors versus chemical biomarkers that are more directly impacted by the targeted gene/protein. This is almost certainly why our p-values for the changes in disease incidence were much larger than our p-values for the biomarkers. No genetic or environmental factor can ever fully account for a complex multifactorial trait like IHD. Additional support for these conclusions can be found in recent studies of recessive disease variants that demonstrated subtle clinical manifestations in heterozygous carriers^51–55^.

While larger and more comprehensive biobanks of linked genomic-phenotypic data can only enhance the functionality of this approach, we have shown that, even with currently available resources like the UK Biobank, it is possible to rely on heterozygous pLOF variants to mimic drug action and thereby predict clinical trial outcomes. One knockout is all you need. The future we have been waiting for – drug targets firmly grounded in human genetics – is already here.

## Data Availability

All data produced in the present work are contained in the manuscript

## Acknowledgements

GKSW initiated the study and, in conjunction with ALS, oversaw its completion. AD performed the computational analyses. GKSW and AD wrote the manuscript. All authors read and revised the manuscript. GKSW is indebted to Maynard Olson for a quarter-century of discussions on the use of human genetics in drug development, and to Lucas DiLeo for creating the title of this manuscript. Funding for this study was provided to ALS through the Natural Sciences and Engineering Research Council of Canada (NSERC) and the Canadian Research Chairs (CRC) program.

## References

1. Paul, S. M. et al. How to improve R&D productivity: the pharmaceutical industry’s grand challenge. Nat. Rev. Drug Discov. 9, 203–214 (2010).

2. DiMasi, J. A., Grabowski, H. G. & Hansen, R. W. Innovation in the pharmaceutical industry: New estimates of R&D costs. J. Health Econ. 47, 20–33 (2016).

3. Arrowsmith, J. & Miller, P. Phase II and Phase III attrition rates 2011–2012. Nat. Rev. Drug Discov. 12, 569–569 (2013).

4. Harrison, R. K. Phase II and phase III failures: 2013–2015. Nat. Rev. Drug Discov. 15, 817–818 (2016).

5. Kola, I. & Landis, J. Can the pharmaceutical industry reduce attrition rates? Nat. Rev. Drug Discov. 3, 711–716 (2004).

6. Waring, M. J. et al. An analysis of the attrition of drug candidates from four major pharmaceutical companies. Nat. Rev. Drug Discov. 14, 475–486 (2015).

7. Hay, M., Thomas, D. W., Craighead, J. L., Economides, C. & Rosenthal, J. Clinical development success rates for investigational drugs. Nat. Biotechnol. 32, 40–51 (2014).

8. McGonigle, P. & Ruggeri, B. Animal models of human disease: Challenges in enabling translation. Biochem. Pharmacol. 87, 162–171 (2014).

9. Horvath, P. et al. Screening out irrelevant cell-based models of disease. Nat. Rev. Drug Discov. 15, 751–769 (2016).

10. Plenge, R. M., Scolnick, E. M. & Altshuler, D. Validating therapeutic targets through human genetics. Nat. Rev. Drug Discov. 12, 581–594 (2013).

11. Nguyen, P. A., Born, D. A., Deaton, A. M., Nioi, P. & Ward, L. D. Phenotypes associated with genes encoding drug targets are predictive of clinical trial side effects. Nat. Commun. 10, 1579 (2019).

12. Nelson, M. R. et al. The support of human genetic evidence for approved drug indications. Nat. Genet. 47, 856–860 (2015).

13. Diogo, D. et al. Phenome-wide association studies across large population cohorts support drug target validation. Nat. Commun. 9, 4285 (2018).

14. King, E. A., Davis, J. W. & Degner, J. F. Are drug targets with genetic support twice as likely to be approved? Revised estimates of the impact of genetic support for drug mechanisms on the probability of drug approval. PLOS Genet. 15, e1008489 (2019).

15. Minikel, E. V., Painter, J. L., Dong, C. C. & Nelson, M. R. Refining the impact of genetic evidence on clinical success. Nature (2024) doi:10.1038/s41586-024-07316-0.

16. Chung, B. H. Y., Chau, J. F. T. & Wong, G. K.-S. Rare versus common diseases: a false dichotomy in precision medicine. NPJ Genomic Med. 6, 19 (2021).

17. MacArthur, D. G. et al. A systematic survey of loss-of-function variants in human protein-coding genes. Science 335, 823–828 (2012).

18. Minikel, E. V. et al. Evaluating drug targets through human loss-of-function genetic variation. Nature 581, 459–464 (2020).

19. Van Hout, C. V. et al. Exome sequencing and characterization of 49,960 individuals in the UK Biobank. Nature 586, 749–756 (2020).

20. Backman, J. D. et al. Exome sequencing and analysis of 454,787 UK Biobank participants. Nature 599, 628–634 (2021).

21. Zschocke, J., Byers, P. H. & Wilkie, A. O. M. Mendelian inheritance revisited: dominance and recessiveness in medical genetics. Nat. Rev. Genet. 24, 442–463 (2023).

22. Morgan, B. & Bridges, A. Sturtevant, The Genetics of Drosophila. Bibliographia Genetica, II. (1925).

23. Orr, H. A. A test of Fisher’s theory of dominance. Proc. Natl. Acad. Sci. 88, 11413–11415 (1991).

24. Deutschbauer, A. M. et al. Mechanisms of Haploinsufficiency Revealed by Genome-Wide Profiling in Yeast. Genetics 169, 1915–1925 (2005).

25. Fisher, R. A. The Possible Modification of the Response of the Wild Type to Recurrent Mutations. Am. Nat. 62, 115–126 (1928).

26. Wright, S. Physiological and Evolutionary Theories of Dominance. Am. Nat. 68, 24–53 (1934).

27. Saleheen, D. et al. Human knockouts and phenotypic analysis in a cohort with a high rate of consanguinity. Nature 544, 235–239 (2017).

28. Lim, E. T. et al. Distribution and Medical Impact of Loss-of-Function Variants in the Finnish Founder Population. PLOS Genet. 10, e1004494 (2014).

29. Gyftopoulos, A. et al. Identification of Novel Genetic Variants and Comorbidities Associated With ICD-10-Based Diagnosis of Hypertrophic Cardiomyopathy Using the UK Biobank Cohort. Front. Genet. 13, 866042 (2022).

30. Ghouse, J., Ahlberg, G., Bundgaard, H. & Olesen, M. S. Effect of Loss-of-Function Genetic Variants in PCSK9 on Glycemic Traits, Neurocognitive Impairment, and Hepatobiliary Function. Diabetes Care 45, 251–254 (2022).

31. Cingolani, P. et al. A program for annotating and predicting the effects of single nucleotide polymorphisms, SnpEff. Fly (Austin) 6, 80–92 (2012).

32. MacArthur, D. G. & Tyler-Smith, C. Loss-of-function variants in the genomes of healthy humans. Hum. Mol. Genet. 19, R125–R130 (2010).

33. Zhan, X., Hu, Y., Li, B., Abecasis, G. R. & Liu, D. J. RVTESTS: an efficient and comprehensive tool for rare variant association analysis using sequence data. Bioinforma. Oxf. Engl. 32, 1423–1426 (2016).

34. Shankar, R. G., Jorgensen, A. & Morris, A. A review of software tools for statistical tests of genetic association with rare variants using next generation sequence data. (2024) doi:10.31219/osf.io/g83ed.

35. Sabatine, M. S. et al. Evolocumab and Clinical Outcomes in Patients with Cardiovascular Disease. N. Engl. J. Med. 376, 1713–1722 (2017).

36. Schwartz, G. G. et al. Alirocumab and Cardiovascular Outcomes after Acute Coronary Syndrome. N. Engl. J. Med. 379, 2097–2107 (2018).

37. Ray, K. K. et al. Two Phase 3 Trials of Inclisiran in Patients with Elevated LDL Cholesterol. N. Engl. J. Med. 382, 1507–1519 (2020).

38. Gouni-Berthold, I. et al. Efficacy and safety of volanesorsen in patients with multifactorial chylomicronaemia (COMPASS): a multicentre, double-blind, randomised, placebo-controlled, phase 3 trial. Lancet Diabetes Endocrinol. 9, 264–275 (2021).

39. Schwabe, C. et al. RNA interference targeting apolipoprotein C-III with ARO-APOC3 in healthy volunteers mimics lipid and lipoprotein findings seen in subjects with inherited apolipoprotein C-III deficiency. Eur. Heart J. 41, (2020).

40. Alexander, V. J. et al. N-acetyl galactosamine-conjugated antisense drug to APOC3 mRNA, triglycerides and atherogenic lipoprotein levels. Eur. Heart J. 40, 2785–2796 (2019).

41. Bergmark, B. A. et al. Effect of Vupanorsen on Non-High-Density Lipoprotein Cholesterol Levels in Statin-Treated Patients With Elevated Cholesterol: TRANSLATE-TIMI 70. Circulation 145, 1377–1386 (2022).

42. Harada-Shiba, M. et al. A randomized study investigating the safety, tolerability, and pharmacokinetics of evinacumab, an ANGPTL3 inhibitor, in healthy Japanese and Caucasian subjects. Atherosclerosis 314, 33–40 (2020).

43. Nissen, S. E. et al. Lepodisiran, an Extended-Duration Short Interfering RNA Targeting Lipoprotein(a): A Randomized Dose-Ascending Clinical Trial. JAMA 330, 2075–2083 (2023).

44. Tsimikas, S. et al. Antisense therapy targeting apolipoprotein(a): a randomised, double-blind, placebo-controlled phase 1 study. Lancet Lond. Engl. 386, 1472–1483 (2015).

45. Koren, M. J. et al. Preclinical development and phase 1 trial of a novel siRNA targeting lipoprotein(a). Nat. Med. 28, 96–103 (2022).

46. Janiszewski, M. et al. A randomized, placebo-controlled, double-blind, ascending single-dose, phase 1 study to evaluate the safety, tolerability, pharmacokinetics, and pharmacodynamics of AMG 529, a novel anti-asgr1 monoclonal antibody, in healthy subjects. J. Am. Coll. Cardiol. 73, 1755–1755 (2019).

47. Kronenberg, F. Human Genetics and the Causal Role of Lipoprotein(a) for Various Diseases. Cardiovasc. Drugs Ther. 30, 87–100 (2016).

48. All of Us Research Program Genomics Investigators. Genomic data in the All of Us Research Program. Nature (2024) doi:10.1038/s41586-023-06957-x.

49. Sullivan, G. M. & Feinn, R. Using Effect Size—or Why the P Value Is Not Enough. J. Grad. Med. Educ. 4, 279–282 (2012).

50. Chen, S. et al. A genomic mutational constraint map using variation in 76,156 human genomes. Nature 625, 92–100 (2024).

51. Barton, A. R., Hujoel, M. L. A., Mukamel, R. E., Sherman, M. A. & Loh, P.-R. A spectrum of recessiveness among Mendelian disease variants in UK Biobank. Am. J. Hum. Genet. 109, 1298–1307 (2022).

52. Nomura, A. et al. Heterozygous ABCG5 Gene Deficiency and Risk of Coronary Artery Disease. Circ. Genomic Precis. Med. 13, 417–423 (2020).

53. Wright, C. F. et al. Assessing the Pathogenicity, Penetrance, and Expressivity of Putative Disease-Causing Variants in a Population Setting. Am. J. Hum. Genet. 104, 275–286 (2019).

54. Çolak, Y., Nordestgaard, B. G. & Afzal, S. Morbidity and mortality in carriers of the cystic fibrosis mutation CFTR Phe508del in the general population. Eur. Respir. J. 56, (2020).

55. Perez, Y. et al. A Rare Variant in PGAP2 Causes Autosomal Recessive Hyperphosphatasia with Mental Retardation Syndrome, with a Mild Phenotype in Heterozygous Carriers. BioMed Res. Int. 2017, 1–7 (2017).

