## Supplementary Tables 1-12 for "Minimizing biological risk for novel inhibitory drug targets: One knockout is all you need"

*^a^ School of Computing, Queen’s University, Kingston, Ontario, Canada, K7L 3N6; ^b^ Department of Molecular and Biomedical Sciences, Queen’s University, Kingston, Ontario, Canada, K7L 3N6; ^c^ Division of Gastroenterology, Department of Medicine, Faculty of Medicine and Dentistry, University of Alberta, Edmonton, Alberta, Canada, T6G 2E1; ^d^ Department of Biological Sciences, Faculty of Science, University of Alberta, Edmonton, Alberta, Canada, T6G 2E1.*

**Supplementary Table 1 | Per gene associations between predicted loss-of-function variants and the complete set of blood biomarkers in the UK Biobank. We also computed the incidence of Ischemic Heart Disease for carriers versus noncarriers.**

| **Variable** | **Noncarriers** | | **Carriers** | **P-Value** | **Change** |
| --- | --- | --- | --- | --- | --- |
| **Sub-table A: *PCSK9* LOF Caucasian Population** | | | | | |
| Mutation Status | | 336,594 | 489 |  |  |
| Age (yr) | | 56.88 ± 7.99 | 56.83 ± 8.34 |  |  |
| Total Cholesterol (mmol/L) | | 5.72 ± 1.14 | 4.94 ± 1.09 | 5.20x10^−53^ | -13.48% |
| LDL Cholesterol (mmol/L) | | 3.57 ± 0.87 | 2.87 ± 0.8 | 4.60x10^−70^ | -19.61% |
| HDL Cholesterol (mmol/L) | | 1.45 ± 0.38 | 1.53 ± 0.43 | 5.01x10^−4^ | +5.16% |
| Remnant Cholesterol (mmol/L) | | 0.69 ± 0.3 | 0.55 ± 0.26 | 2.02x10^−22^ | -20.53% |
| Triglycerides (mmol/L) | | 1.76 ± 1.02 | 1.59 ± 0.93 | 1.95x10^−4^ | -9.38% |
| Lipoprotein A (nmol/L) | | 44.15 ± 49.5 | 43.48 ± 48.23 | 0.81 | -1.53% |
| Alanine aminotransferase (U/L) | | 23.52 ± 13.99 | 22.8 ± 12.9 | 0.77 | -3.05% |
| Albumin (g/L) | | 45.24 ± 2.61 | 45.02 ± 2.73 | 0.12 | -0.49% |
| Alkaline phosphatase (U/L) | | 83.6 ± 26.48 | 83.13 ± 23.18 | 0.61 | -0.55% |
| Apolipoprotein A (g/L) | | 1.54 ± 0.27 | 1.6 ± 0.28 | 2.50x10^−5^ | +3.96% |
| Apolipoprotein B (g/L) | | 1.04 ± 0.24 | 0.85 ± 0.21 | 5.50x10^-66^ | -18.33% |
| Aspartate aminotransferase (U/L) | | 26.19 ± 10.59 | 26.19 ± 10.0 | 0.58 | +0.02% |
| C-reactive protein (mg/L) | | 2.59 ± 4.38 | 2.6 ± 4.34 | 0.65 | +0.49% |
| Calcium (mmol/L) | | 2.38 ± 0.09 | 2.37 ± 0.1 | 3.91x10^−3^ | -0.51% |
| Creatinine (umol/L) | | 72.38 ± 17.81 | 72.02 ± 22.85 | 0.33 | -0.50% |
| Cystatin C (mg/L) | | 0.91 ± 0.17 | 0.91 ± 0.29 | 0.49 | +0.09% |
| Direct bilirubin (umol/L) | | 1.83 ± 0.85 | 1.92 ± 0.93 | 2.44x10^−3^ | +4.48% |
| Gamma glutamyltransferase (U/L) | | 37.33 ± 41.78 | 37.86 ± 40.93 | 0.57 | +1.41% |
| Glucose (mmol/L) | | 5.12 ± 1.21 | 5.09 ± 1.11 | 0.76 | -0.47% |
| Glycated haemoglobin (HbA1c) (mmol/mol) | | 35.95 ± 6.51 | 35.66 ± 5.85 | 0.39 | -0.83% |
| IGF-1 (nmol/L) | | 21.41 ± 5.66 | 21.39 ± 5.7 | 0.91 | -0.10% |
| Oestradiol (pmol/L) | | 459.18 ± 436.21 | 542.31 ± 511.62 | 0.03 | +18.10% |
| Phosphate (mmol/L) | | 1.16 ± 0.16 | 1.15 ± 0.14 | 0.11 | -0.66% |
| Rheumatoid factor (IU/ml) | | 24.46 ± 19.67 | 24.03 ± 21.37 | 0.57 | -1.74% |
| SHBG (nmol/L) | | 51.82 ± 27.65 | 54.52 ± 26.64 | 0.20 | +5.22% |
| Testosterone (nmol/L) | | 6.61 ± 6.05 | 6.14 ± 6.2 | 0.35 | -7.18% |
| Total bilirubin (umol/L) | | 9.15 ± 4.43 | 8.92 ± 4.12 | 0.74 | -2.51% |
| Total protein (g/L) | | 72.35 ± 4.03 | 71.92 ± 4.23 | 0.03 | -0.60% |
| Urate (umol/L) | | 309.51 ± 80.3 | 298.95 ± 74.0 | 0.11 | -3.41% |
| Urea (mmol/L) | | 5.44 ± 1.39 | 5.39 ± 1.43 | 0.68 | -0.83% |
| Vitamin D (nmol/L) | | 49.82 ± 20.95 | 51.32 ± 21.61 | 0.14 | +3.02% |
| Ischemic heart disease – no. subjects | | 39,699 (11.8%) | 40 (8.2%) | 0.02 | -30.65% |
| **Sub-table B: *APOC3* LOF Caucasian Population** | | | | | |
| Mutation Status | | 335,446 | 1,637 |  |  |
| Age (yr) | | 56.88 ± 7.99 | 56.88 ± 7.74 |  |  |
| Total Cholesterol (mmol/L) | | 5.71±1.14 | 5.69±1.04 | 0.69 | -0.39% |
| LDL Cholesterol (mmol/L) | | 3.57 ± 0.87 | 3.42 ± 0.77 | 9.32x10^−12^ | -4.34% |
| HDL Cholesterol (mmol/L) | | 1.45 ± 0.38 | 1.78 ± 0.43 | 8.04x10^-283^ | +22.39% |
| Remnant Cholesterol (mmol/L) | | 0.69 ± 0.3 | 0.49 ± 0.2 | 3.37x10^−137^ | -28.43% |
| Triglycerides (mmol/L) | | 1.76 ± 1.02 | 0.93 ± 0.43 | 1.10x10^−236^ | -46.95% |
| Lipoprotein A (nmol/L) | | 44.15 ± 49.5 | 43.75 ± 48.97 | 0.61 | -0.92% |
| Alanine aminotransferase (U/L) | | 23.52 ± 13.99 | 24.16 ± 14.17 | 0.38 | +2.73% |
| Albumin (g/L) | | 45.24 ± 2.61 | 45.54 ± 2.71 | 1.34x10^−5^ | +0.66% |
| Alkaline phosphatase (U/L) | | 83.59 ± 26.48 | 85.03 ± 24.95 | 0.03 | +1.73% |
| Apolipoprotein A (g/L) | | 1.54 ± 0.27 | 1.73 ± 0.29 | 2.02x10^−193^ | +12.48% |
| Apolipoprotein B (g/L) | | 1.04 ± 0.24 | 0.99 ± 0.21 | 2.91x10^−14^ | -4.49% |
| Aspartate aminotransferase (U/L) | | 26.19 ± 10.6 | 25.97 ± 8.94 | 0.24 | -0.85% |
| C-reactive protein (mg/L) | | 2.58 ± 4.37 | 3.01 ± 4.75 | 1.95x10^−7^ | +16.69% |
| Calcium (mmol/L) | | 2.38 ± 0.09 | 2.39 ± 0.09 | 1.67x10^−3^ | +0.30% |
| Creatinine (umol/L) | | 72.37 ± 17.83 | 73.7 ± 15.16 | 0.01 | +1.83% |
| Cystatin C (mg/L) | | 0.91 ± 0.17 | 0.92 ± 0.15 | 0.04 | +1.12% |
| Direct bilirubin (umol/L) | | 1.83 ± 0.86 | 1.88 ± 0.81 | 0.02 | +2.57% |
| Gamma glutamyltransferase (U/L) | | 37.33 ± 41.77 | 38.67 ± 42.43 | 0.12 | +3.59% |
| Glucose (mmol/L) | | 5.12 ± 1.21 | 5.13 ± 1.22 | 0.71 | +0.26% |
| Glycated haemoglobin (mmol/mol) | | 35.95 ± 6.5 | 36.4 ± 6.87 | 7.36x10^−3^ | +1.24% |
| IGF-1 (nmol/L) | | 21.41 ± 5.66 | 21.49 ± 5.55 | 0.49 | +0.39% |
| Oestradiol (pmol/L) | | 459.44 ± 436.84 | 436.38 ± 327.09 | 0.76 | -5.02% |
| Phosphate (mmol/L) | | 1.16 ± 0.16 | 1.16 ± 0.17 | 0.21 | +0.27% |
| Rheumatoid factor (IU/ml) | | 24.46 ± 19.68 | 23.94 ± 19.32 | 0.76 | -2.13% |
| SHBG (nmol/L) | | 51.83 ± 27.64 | 50.47 ± 28.18 | 0.17 | -2.61% |
| Testosterone (nmol/L) | | 6.61 ± 6.05 | 6.88 ± 6.04 | 0.61 | +4.14% |
| Total bilirubin (umol/L) | | 9.15 ± 4.43 | 9.02 ± 4.27 | 0.04 | -1.40% |
| Total protein (g/L) | | 72.35 ± 4.03 | 72.77 ± 4.17 | 9.30x10^−5^ | +0.58% |
| Urate (umol/L) | | 309.48 ± 80.3 | 313.11 ± 79.49 | 0.38 | +1.17% |
| Urea (mmol/L) | | 5.43 ± 1.39 | 5.54 ± 1.36 | 3.38x10^−3^ | +1.92% |
| Vitamin D (nmol/L) | | 49.82 ± 20.96 | 50.12 ± 20.45 | 0.37 | +0.61% |
| Ischemic heart disease – no. subjects | | 39,572 (11.8%) | 167 (10.2%) | 0.02 | -14.04% |
| **Sub-table C: *ANGPTL3* LOF Caucasian Population** | | | | | |
| Mutation Status | | 336,460 | 623 |  |  |
| Age (yr) | | 56.88 ± 7.99 | 56.69 ± 7.99 |  |  |
| Total Cholesterol (mmol/L) | | 5.72 ± 1.14 | 5.1 ± 0.99 | 6.81x10^−41^ | -10.72% |
| LDL Cholesterol (mmol/L) | | 3.57 ± 0.87 | 3.23 ± 0.77 | 1.54x10^−21^ | -9.48% |
| HDL Cholesterol (mmol/L) | | 1.45 ± 0.38 | 1.34 ± 0.36 | 1.06x10^-14^ | -7.75% |
| Remnant Cholesterol (mmol/L) | | 0.69 ± 0.3 | 0.53 ± 0.22 | 1.58x10^−36^ | -23.56% |
| Triglycerides (mmol/L) | | 1.76 ± 1.02 | 1.22 ± 0.6 | 7.43x10^−57^ | -30.62% |
| Lipoprotein A (nmol/L) | | 44.16 ± 49.5 | 40.93 ± 46.83 | 0.18 | -7.30% |
| Alanine aminotransferase (U/L) | | 23.52 ± 13.98 | 23.7 ± 18.47 | 0.64 | +0.76% |
| Albumin (g/L) | | 45.24 ± 2.61 | 45.33 ± 2.51 | 0.44 | +0.19% |
| Alkaline phosphatase (U/L) | | 83.6 ± 26.48 | 82.37 ± 23.48 | 0.28 | -1.47% |
| Apolipoprotein A (g/L) | | 1.54 ± 0.27 | 1.41 ± 0.25 | 7.00x10^−39^ | -8.89% |
| Apolipoprotein B (g/L) | | 1.03 ± 0.24 | 0.98 ± 0.21 | 3.49x10^−9^ | -5.58% |
| Aspartate aminotransferase (U/L) | | 26.19 ± 10.59 | 25.42 ± 9.39 | 0.09 | 2.94% |
| C-reactive protein (mg/L) | | 2.59 ± 4.38 | 2.41 ± 4.16 | 1.67x10^−3^ | -6.86% |
| Calcium (mmol/L) | | 2.38 ± 0.09 | 2.37 ± 0.09 | 0.03 | -0.36% |
| Creatinine (umol/L) | | 72.38 ± 17.82 | 71.36 ± 15.35 | 0.24 | -1.41% |
| Cystatin C (mg/L) | | 0.91 ± 0.17 | 0.9 ± 0.17 | 0.58 | -0.68% |
| Direct bilirubin (umol/L) | | 1.83 ± 0.86 | 1.84 ± 0.85 | 0.95 | +0.04% |
| Gamma glutamyltransferase (U/L) | | 37.34 ± 41.79 | 33.64 ± 30.61 | 0.05 | -9.93% |
| Glucose (mmol/L) | | 5.12 ± 1.21 | 5.17 ± 1.39 | 0.24 | +1.06% |
| Glycated haemoglobin (mmol/mol) | | 35.95 ± 6.5 | 36.26 ± 7.6 | 0.17 | +0.86% |
| IGF-1 (nmol/L) | | 21.41 ± 5.66 | 21.34 ± 5.64 | 0.66 | -0.34% |
| Oestradiol (pmol/L) | | 459.33 ± 436.55 | 456.24 ± 332.46 | 0.88 | -0.67% |
| Phosphate (mmol/L) | | 1.16 ± 0.16 | 1.16 ± 0.16 | 0.81 | -0.07% |
| Rheumatoid factor (IU/ml) | | 24.46 ± 19.68 | 23.61 ± 18.3 | 0.82 | -3.47% |
| SHBG (nmol/L) | | 51.82 ± 27.65 | 51.75 ± 26.17 | 0.78 | -0.14% |
| Testosterone (nmol/L) | | 6.61 ± 6.05 | 6.72 ± 6.24 | 0.25 | +1.72% |
| Total bilirubin (umol/L) | | 9.15 ± 4.43 | 8.81 ± 4.18 | 0.03 | -3.67% |
| Total protein (g/L) | | 72.35 ± 4.03 | 72.12 ± 3.75 | 0.18 | -0.32% |
| Urate (umol/L) | | 309.5 ± 80.29 | 307.48 ± 79.48 | 0.79 | -0.66% |
| Urea (mmol/L) | | 5.44 ± 1.39 | 5.44 ± 1.47 | 0.68 | +0.12% |
| Vitamin D (nmol/L) | | 49.81 ± 20.95 | 53.31 ± 22.33 | 8.37x10^-5^ | +6.60% |
| Ischemic heart disease – no. subjects | | 39,680 (11.8%) | 59 (9.5%) | 0.11 | -19.70% |
| **Sub-table D: *LPA* LOF Caucasian Population** | | | | | |
| Mutation Status | | 310,336 | 26,747 |  |  |
| Age (yr) | | 56.88 ± 7.99 | 56.89 ± 7.99 |  |  |
| Total Cholesterol (mmol/L) | | 5.72 ± 1.14 | 5.71 ± 1.13 | 0.20 | -0.17% |
| LDL Cholesterol (mmol/L) | | 3.57 ± 0.87 | 3.56 ± 0.86 | 3.73x10^−3^ | -0.47% |
| HDL Cholesterol (mmol/L) | | 1.45 ± 0.38 | 1.46 ± 0.39 | 6.48x10^−5^ | +0.63% |
| Remnant Cholesterol (mmol/L) | | 0.69 ± 0.3 | 0.69 ± 0.3 | 0.38 | -0.25% |
| Triglycerides (mmol/L) | | 1.75 ± 1.02 | 1.76 ± 1.03 | 0.36 | +0.39% |
| Lipoprotein A (nmol/L) | | 44.82 ± 49.68 | 33.62 ± 45.13 | 2.42x10^−164^ | -24.99% |
| Alanine aminotransferase (U/L) | | 23.52 ± 13.92 | 23.58 ± 14.76 | 0.48 | +0.29% |
| Albumin (g/L) | | 45.24 ± 2.61 | 45.26 ± 2.62 | 0.15 | +0.05% |
| Alkaline phosphatase (U/L) | | 83.6 ± 26.43 | 83.56 ± 26.98 | 0.77 | -0.05% |
| Apolipoprotein A (g/L) | | 1.54 ± 0.27 | 1.55 ± 0.27 | 0.01 | +0.25% |
| Apolipoprotein B (g/L) | | 1.04 ± 0.24 | 1.03 ± 0.24 | 1.20x10^−3^ | -0.57% |
| Aspartate aminotransferase (U/L) | | 26.19 ± 10.56 | 26.19 ± 10.94 | 0.97 | -0.02% |
| C-reactive protein (mg/L) | | 2.59 ± 4.38 | 2.58 ± 4.34 | 0.68 | -0.25% |
| Calcium (mmol/L) | | 2.38 ± 0.09 | 2.38 ± 0.09 | 0.09 | +0.04% |
| Creatinine (umol/L) | | 72.37 ± 17.75 | 72.47 ± 18.6 | 0.43 | +0.13% |
| Cystatin C (mg/L) | | 0.91 ± 0.17 | 0.91 ± 0.17 | 0.15 | -0.14% |
| Direct bilirubin (umol/L) | | 1.83 ± 0.85 | 1.84 ± 0.89 | 0.69 | +0.09% |
| Gamma glutamyltransferase (U/L) | | 37.33 ± 41.69 | 37.4 ± 42.8 | 0.94 | +0.19% |
| Glucose (mmol/L) | | 5.12 ± 1.2 | 5.13 ± 1.23 | 0.26 | +0.17% |
| Glycated haemoglobin (mmol/mol) | | 35.95 ± 6.52 | 35.96 ± 6.31 | 0.97 | +0.01% |
| IGF-1 (nmol/L) | | 21.4 ± 5.66 | 21.47 ± 5.66 | 0.03 | +0.32% |
| Oestradiol (pmol/L) | | 459.02 ± 431.66 | 462.87 ± 487.13 | 0.47 | +0.84% |
| Phosphate (mmol/L) | | 1.16 ± 0.16 | 1.16 ± 0.16 | 0.42 | -0.07% |
| Rheumatoid factor (IU/ml) | | 24.47 ± 19.7 | 24.26 ± 19.37 | 0.48 | -0.86% |
| SHBG (nmol/L) | | 51.82 ± 27.63 | 51.8 ± 27.84 | 0.96 | -0.05% |
| Testosterone (nmol/L) | | 6.61 ± 6.05 | 6.6 ± 6.05 | 0.67 | -0.19% |
| Total bilirubin (umol/L) | | 9.15 ± 4.42 | 9.13 ± 4.52 | 0.22 | -0.16% |
| Total protein (g/L) | | 72.35 ± 4.03 | 72.36 ± 4.04 | 0.76 | +0.01% |
| Urate (umol/L) | | 309.55 ± 80.26 | 308.92 ± 80.65 | 0.07 | -0.20% |
| Urea (mmol/L) | | 5.44 ± 1.39 | 5.43 ± 1.37 | 0.67 | -0.06% |
| Vitamin D (nmol/L) | | 49.84 ± 20.97 | 49.62 ± 20.83 | 0.28 | -0.43% |
| Ischemic heart disease – no. subjects | | 36,733 | 3,006 | 9.71x10^−4^ | -5.05% |
| **Sub-table E: *ASGR1* LOF Caucasian Population** | | | | | |
| Mutation Status | | 336,962 | 121 |  |  |
| Age (yr) | | 56.88 ± 7.99 | 56.87 ± 8.07 |  |  |
| Total Cholesterol (mmol/L) | | 5.71 ± 1.14 | 5.45 ± 1.08 | 0.02 | -4.60% |
| LDL Cholesterol (mmol/L) | | 3.57 ± 0.87 | 3.37 ± 0.78 | 0.02 | -5.65% |
| HDL Cholesterol (mmol/L) | | 1.45 ± 0.38 | 1.47 ± 0.41 | 0.41 | +1.05% |
| Remnant Cholesterol (mmol/L) | | 0.69 ± 0.3 | 0.63 ± 0.3 | 0.03 | -8.96% |
| Triglycerides (mmol/L) | | 1.76 ± 1.02 | 1.73 ± 1.02 | 0.41 | -1.61% |
| Lipoprotein A (nmol/L) | | 44.15 ± 49.49 | 38.71 ± 47.41 | 0.20 | -12.32% |
| Alanine aminotransferase (U/L) | | 23.52 ± 13.99 | 27.02 ± 19.03 | 0.04 | +14.88% |
| Albumin (g/L) | | 45.24 ± 2.61 | 45.11 ± 2.5 | 0.45 | -0.28% |
| Alkaline phosphatase (U/L) | | 83.59 ± 26.46 | 110.91 ± 33.36 | 1.87x10^−30^ | +32.69% |
| Apolipoprotein A (g/L) | | 1.54 ± 0.27 | 1.53 ± 0.29 | 0.77 | -0.98% |
| Apolipoprotein B (g/L) | | 1.03 ± 0.24 | 0.98 ± 0.23 | 0.01 | -5.56% |
| Aspartate aminotransferase (U/L) | | 26.19 ± 10.59 | 27.92 ± 13.81 | 0.10 | +6.62% |
| C-reactive protein (mg/L) | | 2.59 ± 4.38 | 2.5 ± 3.9 | 0.94 | -3.14% |
| Calcium (mmol/L) | | 2.38 ± 0.09 | 2.38 ± 0.08 | 0.90 | +0.04% |
| Creatinine (umol/L) | | 72.38 ± 17.82 | 72.46 ± 12.75 | 0.50 | +0.11% |
| Cystatin C (mg/L) | | 0.91 ± 0.17 | 0.89 ± 0.13 | 0.18 | -1.94% |
| Direct bilirubin (umol/L) | | 1.83 ± 0.86 | 1.93 ± 0.89 | 0.24 | +5.39% |
| Gamma Glutamyltransferase (U/L) | | 37.33 ± 41.74 | 51.18 ± 98.11 | 0.03 | +37.09% |
| Glucose (mmol/L) | | 5.12 ± 1.21 | 5.09 ± 0.76 | 0.85 | -0.51% |
| Glycated haemoglobin (mmol/mol) | | 35.95 ± 6.51 | 36.54 ± 7.31 | 0.35 | +1.62% |
| IGF-1 (nmol/L) | | 21.41 ± 5.66 | 21.23 ± 5.7 | 0.54 | -0.84% |
| Oestradiol (pmol/L) | | 459.38 ± 436.44 | 328.48 ± 163.46 | 0.48 | -28.49% |
| Phosphate (mmol/L) | | 1.16 ± 0.16 | 1.17 ± 0.16 | 0.31 | +1.12% |
| Rheumatoid factor (IU/ml) | | 24.46 ± 19.68 | 17.4 ± 5.06 | 0.49 | -28.86% |
| SHBG (nmol/L) | | 51.82 ± 27.64 | 47.64 ± 26.25 | 0.16 | -8.08% |
| Testosterone (nmol/L) | | 6.61 ± 6.05 | 6.71 ± 5.52 | 0.02 | +1.56% |
| Total bilirubin (umol/L) | | 9.15 ± 4.43 | 9.55 ± 4.62 | 0.49 | +4.44% |
| Total protein (g/L) | | 72.35 ± 4.03 | 72.61 ± 3.32 | 0.54 | +0.36% |
| Urate (umol/L) | | 309.5 ± 80.29 | 309.65 ± 83.65 | 0.52 | +0.05% |
| Urea (mmol/L) | | 5.44 ± 1.39 | 5.44 ± 1.31 | 0.92 | +0.10% |
| Vitamin D (nmol/L) | | 49.82 ± 20.95 | 50.69 ± 22.77 | 0.71 | +1.75% |
| Ischemic heart disease – no. subjects | | 39,722 (11.8%) | 17 (14.0%) | 0.49 | +19.18% |

**Supplementary Table 2 | Comparison of our genetic study with other published genetic studies.**

| **Sub-Table A: *PCSK9*** | | | | | | | | | | | | | | | |
| --- | --- | --- | --- | --- | --- | --- | --- | --- | --- | --- | --- | --- | --- | --- | --- |
|  | Our Study | | | Ghouse *et al.* | | | | Cohen *et al.* | | | | |  | | |
|  | Noncarriers | Carriers | | Noncarriers | | Carriers | | Noncarriers | | Carriers | | |  | |  |
|  | 406,035 | 620 | | 166,742 | | 374 | | 9,223 | | 301 | | |  | |  |
| Variable | Change | P-Value | | Change | | P-Value | | Change | | P-Value | | |  | |  |
| Total Cholesterol (mmol/L) | -13% | 5.20x10^−53^ | | - | | - | | -9% | | < 0.001 | | |  | |  |
| LDL Cholesterol (mmol/L) | -19% | 4.60x10^−70^ | | -19% | | 4.60x10^-55^ | | -15% | | < 0.001 | | |  | |  |
| HDL Cholesterol (mmol/L) | +5% | 5.01x10^-4^ | | +0.7% | | 0.28 | | +2% | | 0.64 | | |  | |  |
| Remnant Cholesterol (mmol/L) | -20% | 2.02x10^-28^ | | - | | - | | -18% | | N/A | | |  | |  |
| Triglycerides (mmol/L) | -9% | 1.95x10^-4^ | | -7% | | 0.159 | | -16% | | 0.04 | | |  | |  |
| Apolipoprotein A (g/L) | +3% | 2.50x10^-5^ | | +2% | | 0.016 | | - | | - | | |  | |  |
| Apolipoprotein B (g/L) | -18% | 5.50x10^-66^ | | -18% | | 7.60x10^-50^ | | - | | - | | |  | |  |
| Ischemic heart disease – no. subjects | -30% | 0.02 | | - | | - | | -46% | | 0.003 | | |  | |  |
| **Sub-table B: *APOC3*** | | | | | | | | | | | | | | | |
|  | Our Study | | | | Wulff *et al.* | | | TG and HDL group | | | | Pokharel *et al.* | | | |
|  | Noncarriers | | Carriers | | Noncarriers | | Carriers | Noncarriers | | Carriers | | Noncarriers | | | Carriers |
|  | 406,631 | | 2,024 | | 75,465 | | 260 | 3,701 | | 33 | | 4,501 | | | 23 |
| Variable | Change | | P-Value | | Change | | P-Value | Change | | P-Value | | Change | | | P-Value |
| LDL Cholesterol (mmol/L) | -4% | | 9.32x10^−12^ | | -3% | | 0.06 | -16% | | 0.05 | | -9% | | | 0.04 |
| HDL Cholesterol (mmol/L) | +22% | | 8.04x10^−283^ | | +33% | | 2x10^−29^ | +22% | | 4x10^−6^ | | +36% | | | 1.00x10^−4^ |
| Remnant Cholesterol (mmol/L) | -28% | | 3.37x10^−137^ | | -43% | | 5x10^-49^ | - | | - | | -51% | | | N/A |
| Triglycerides (mmol/L) | -47% | | 1.10x10^-236^ | | -47% | | 3x10^-54^ | -38.5% | | 6x10^-9^ | | -52% | | | 1x10^-4^ |
| Apolipoprotein B (g/L) | -4% | | 2.91x10^-14^ | | -13% | | 4x10^-19^ | - | | - | | -14% | | | 0.49 |
| Ischemic heart disease – no. subjects | -14% | | 0.02 | | -22% | | 0.22 | -40% | | 4x10^-6^ | | - | | | - |
| **Sub-table C: *ANGPTL3*** | | | | | | | | | | | | | | | |
|  | Our Study | | | | Robciuc *et al.* | | | | Dewey *et al.* | | | | | Stitziel *et al.* | |
|  | Noncarriers | | Carriers | | Noncarriers | | Carriers | | Noncarriers | | Carriers | | | Noncarriers | Carriers |
|  | 405,904 | | 751 | | 22 | | 17 | | 45,036 | | 191 | | | 20,092 | 60 |
| Variable | Change | | P-Value | | Change | | P-Value | | Change | | P-Value | | | Change | P-Value |
| Total Cholesterol (mmol/L) | -10% | | 6.81x10^-41^ | | -11% | | N/A | | -12% | | 1.7x10^-17^ | | | -10% | 8x10^-4^ |
| LDL Cholesterol (mmol/L) | -9% | | 1.54x10^-21^ | | -16% | | N/A | | -7% | | 2.8x10^-5^ | | | -11% | 0.04 |
| HDL Cholesterol (mmol/L) | -7% | | 1.06x10^-14^ | | 0% | | N/A | | -6% | | 0.02 | | | -5% | 0.17 |
| Remnant Cholesterol (mmol/L) | -23% | | 1.58x10^−36^ | | -16% | | N/A | | -38% | | N/A | | | - | - |
| Triglycerides (mmol/L) | -30% | | 7.43x10^-57^ | | -26% | | N/A | | -27% | | 5x10^-21^ | | | -17% | 0.01 |
| Ischemic heart disease – no. subjects | -19% | | 0.11 | | - | | - | | -39% | | 4x10^-3^ | | | -34% | 0.04 |
| **Sub-table D: *LPA*** | | | | | | | | | | | | | | | |
|  | Our Study | | | | Kyriakou *et al.* | | | |  | | | | |  | |
|  | Noncarriers | | Carriers | | Noncarriers | | Carriers | |  | |  | | |  |  |
|  | 310,336 | | 26,747 | | 1,764 | | 93 | |  | |  | | |  |  |
| Variable | Change | | P-Value | | Change | | P-Value | |  | |  | | |  |  |
| Lipoprotein A (mmol/L) | -25% | | 2.42x10^-164^ | | -39% | | 2x10^-10^ | |  | |  | | |  |  |
| Ischemic heart disease – no. subjects | -5% | | 9.71x10^-4^ | | -21% | | 0.023 | |  | |  | | |  |  |
| **Sub-table E: *ASGR1*** | | | | | | | | | | | | | | | |
|  | Our Study | | | | Nioi *et al.* | | | |  | | | | |  | |
|  | Noncarriers | | Carriers | | Noncarriers | | Carriers | |  | |  | | |  |  |
|  | 336,962 | | 121 | | 119,514 | | 490 | |  | |  | | |  |  |
| Variable | Change | | P-Value | | Change | | P-Value | |  | |  | | |  |  |
| Total Cholesterol (mmol/L) | -4% | | 0.02 | | -11% | | N/A | |  | |  | | |  |  |
| LDL Cholesterol (mmol/L) | -5% | | 0.02 | | -9% | | 3.9x10^-11^ | |  | |  | | |  |  |
| Alkaline phosphatase (U/L) | +32% | | 1.87x10^-30^ | | +46% | | 5.6x10^-69^ | |  | |  | | |  |  |
| Ischemic heart disease – no. subjects | +19% | | 0.49 | | -34% | | 4.0x10^-6^ | |  | |  | | |  |  |

**Supplementary Table 3 | *PCSK9* Predicted Loss-of-Function Variants. Allele frequency is estimated by UK Biobank.**

| **Chromosome** | **Position** | **Reference Allele** | **Alternate Allele** | **Allele Frequency** | **Allele Quality** | **Variant Type** | **Mutation** |
| --- | --- | --- | --- | --- | --- | --- | --- |
| chr1 | 55039838 | A | G | AF=5e-06 | AQ=48 | start lost | p.Met1? |
| chr1 | 55039840 | G | T | AF=1e-06 | AQ=48 | start lost | p.Met1? |
| chr1 | 55040006 | G | T | AF=6e-06 | AQ=43 | stop gained | p.Glu57* |
| chr1 | 55040024 | AC | A | AF=7e-06 | AQ=42 | frameshift variant | p.Phe64fs |
| chr1 | 55043851 | G | A | AF=1e-06 | AQ=44 | stop gained | p.Trp72* |
| chr1 | 55043903 | C | T | AF=1e-06 | AQ=44 | stop gained | p.Gln90* |
| chr1 | 55043930 | C | T | AF=1e-06 | AQ=44 | stop gained | p.Gln99* |
| chr1 | 55043998 | CTTCCTGGT | C | AF=1e-06 | AQ=48 | frameshift variant | p.Phe122fs |
| chr1 | 55044023 | CTGCTG | C | AF=3e-06 | AQ=33 | frameshift variant | p.Leu130fs |
| chr1 | 55046570 | CT | C | AF=1e-06 | AQ=48 | frameshift variant | p.Phe150fs |
| chr1 | 55046570 | CTT | C | AF=1e-06 | AQ=37 | frameshift variant | p.Phe150fs |
| chr1 | 55046621 | C | A | AF=1e-06 | AQ=37 | stop gained | p.Tyr166* |
| chr1 | 55052273 | GA | G | AF=3e-06 | AQ=47 | splice acceptor variant & c.524-2delA |  |
| chr1 | 55052374 | ATG | A | AF=3e-06 | AQ=40 | frameshift variant | p.Val208fs |
| chr1 | 55052386 | AGGACGGGACCC | A | AF=1.3e-05 | AQ=52 | frameshift variant | p.Asp212fs |
| chr1 | 55052412 | G | T | AF=0.000142 | AQ=53 | splice donor variant & intron variant | c.657+1G*>*T |
| chr1 | 55052413 | T | C | AF=1e-06 | AQ=50 | splice donor variant & intron variant | c.657+2T*>*C |
| chr1 | 55052648 | A | C | AF=2e-06 | AQ=50 | splice acceptor variant & intron variant | c.658-2A*>*C |
| chr1 | 55052704 | G | GA | AF=5e-06 | AQ=59 | frameshift variant | p.Asp238fs |
| chr1 | 55052746 | GT | G | AF=1.1e-05 | AQ=50 | frameshift variant | p.Val252fs |
| chr1 | 55056006 | TC | T | AF=5e-06 | AQ=52 | frameshift variant | p.Arg272fs |
| chr1 | 55056009 | GA | G | AF=1.3e-05 | AQ=52 | frameshift variant | p.Ser274fs |
| chr1 | 55056146 | TC | T | AF=1e-06 | AQ=42 | frameshift variant | p.Arg319fs |
| chr1 | 55057364 | C | T | AF=1.3e-05 | AQ=56 | stop gained | p.Gln344* |
| chr1 | 55057468 | C | A | AF=6e-06 | AQ=44 | stop gained | p.Cys378* |
| chr1 | 55057478 | C | T | AF=2.4e-05 | AQ=49 | stop gained | p.Gln382* |
| chr1 | 55058092 | CA | C | AF=1e-06 | AQ=41 | frameshift variant | p.Gln413fs |
| chr1 | 55058113 | GC | G | AF=3e-05 | AQ=52 | frameshift variant | p.Asp422fs |
| chr1 | 55058139 | G | A | AF=4e-06 | AQ=44 | stop gained | p.Trp428* |
| chr1 | 55058597 | A | AG | AF=1e-06 | AQ=41 | frameshift variant | p.Ser485fs |
| chr1 | 55058649 | T | C | AF=2e-06 | AQ=36 | splice donor variant & intron variant | c.1503+2T*>*C |
| chr1 | 55059491 | A | AG | AF=1e-06 | AQ=43 | frameshift variant | p.Lys506fs |
| chr1 | 55059573 | C | T | AF=1e-06 | AQ=43 | stop gained | p.Gln531* |
| chr1 | 55061368 | G | GC | AF=5.6e-05 | AQ=50 | splice acceptor variant & intron variant | c.1682-3dupC |
| chr1 | 55061374 | G | C | AF=3e-06 | AQ=51 | splice acceptor variant & intron variant | c.1682-1G*>*C |
| chr1 | 55061435 | CA | C | AF=1e-06 | AQ=37 | frameshift variant | p.Arg582fs |
| chr1 | 55061437 | C | T | AF=7e-06 | AQ=43 | stop gained | p.Arg582* |
| chr1 | 55061473 | GC | G | AF=3e-06 | AQ=45 | frameshift variant | p.Ser595fs |
| chr1 | 55061492 | GC | G | AF=2e-06 | AQ=49 | frameshift variant | p.Cys601fs |
| chr1 | 55061557 | G | A | AF=0.000298 | AQ=52 | splice donor variant & intron variant | c.1863+1G*>*A |
| chr1 | 55063433 | AC | A | AF=3e-06 | AQ=47 | frameshift variant | p.His643fs |

**Supplementary Table 4 | *APOC3* Predicted Loss-of-Function Variants. Allele frequency is estimated by UK Biobank.**

| **Chromosome** | **Position** | **Reference Allele** | **Alternate Allele** | **Allele Frequency** | **Allele Quality** | **Variant Type** | **Mutation** |
| --- | --- | --- | --- | --- | --- | --- | --- |
| chr11 | 116830568 | A | G | AF=1e-05 | AQ=45 | splice acceptor variant & intron variant | c.-13-2A*>*G |
| chr11 | 116830584 | T | C | AF=1e-06 | AQ=41 | start lost | p.Met1? |
| chr11 | 116830616 | C | CT | AF=6e-06 | AQ=46 | frameshift variant | p.Ala13fs |
| chr11 | 116830637 | C | T | AF=0.000263 | AQ=49 | stop gained & splice region variant | p.Arg19* |
| chr11 | 116830638 | G | A | AF=0.002204 | AQ=53 | splice donor variant & intron variant | c.55+1G*>*A |
| chr11 | 116830638 | G | T | AF=1.6e-05 | AQ=42 | splice donor variant & intron variant | c.55+1G*>*T |
| chr11 | 116830897 | G | T | AF=3.9e-05 | AQ=44 | splice donor variant & intron variant | c.179+1G*>*T |

**Supplementary Table 5 | *ANGPTL3* Predicted Loss-of-Function Variants. Allele frequency is estimated by UK Biobank.**

| **Chromosome** | **Position** | **Reference Allele** | **Alternate Allele** | **Allele Frequency** | **Allele Quality** | **Variant Type** | **Mutation** |
| --- | --- | --- | --- | --- | --- | --- | --- |
| chr1 | 62597562 | T | TA | AF=2e-06 | AQ=37 | frameshift variant & start lost | p.Met1fs |
| chr1 | 62597634 | A | AT | AF=1e-06 | AQ=44 | frameshift variant | p.Ser24fs |
| chr1 | 62597670 | C | G | AF=1e-06 | AQ=38 | stop gained | p.Ser35* |
| chr1 | 62597731 | TG | T | AF=2e-06 | AQ=49 | frameshift variant | p.Gly56fs |
| chr1 | 62597742 | AC | A | AF=3e-06 | AQ=43 | frameshift variant | p.Phe60fs |
| chr1 | 62597759 | A | T | AF=1e-06 | AQ=42 | stop gained | p.Lys65* |
| chr1 | 62597909 | AAT | A | AF=5e-06 | AQ=51 | frameshift variant | p.Met116fs |
| chr1 | 62597921 | GAACTC | G | AF=0.000577 | AQ=65 | frameshift variant | p.Asn121fs |
| chr1 | 62597954 | G | GA | AF=2e-06 | AQ=36 | frameshift variant | p.Ile132fs |
| chr1 | 62597972 | CAA | C | AF=1e-06 | AQ=55 | frameshift variant | p.Lys137fs |
| chr1 | 62597996 | CAACT | C | AF=0.000204 | AQ=53 | frameshift variant | p.Asn147fs |
| chr1 | 62598020 | C | T | AF=1e-06 | AQ=41 | stop gained | p.Gln152* |
| chr1 | 62598711 | C | T | AF=2e-06 | AQ=46 | stop gained | p.Gln171* |
| chr1 | 62598753 | CA | C | AF=5e-06 | AQ=52 | frameshift variant | p.Gln185fs |
| chr1 | 62598762 | CAATT | C | AF=1e-06 | AQ=39 | frameshift variant | p.Leu189fs |
| chr1 | 62598771 | CA | C | AF=1.2e-05 | AQ=48 | frameshift variant | p.Gln191fs |
| chr1 | 62598774 | C | T | AF=3e-06 | AQ=48 | stop gained | p.Gln192* |
| chr1 | 62601080 | A | G | AF=1e-06 | AQ=39 | splice acceptor variant & intron variant | c.607-2A*>*G |
| chr1 | 62601115 | AT | A | AF=2.8e-05 | AQ=53 | frameshift variant | p.Ser215fs |
| chr1 | 62601870 | GATGTT | G | AF=1e-06 | AQ=35 | frameshift variant | p.Val276fs |
| chr1 | 62601880 | C | A | AF=1.4e-05 | AQ=44 | stop gained & splice region variant | p.Ser278* |
| chr1 | 62601880 | C | G | AF=5e-06 | AQ=48 | stop gained & splice region variant | p.Ser278* |
| chr1 | 62602311 | C | T | AF=6e-06 | AQ=54 | stop gained | p.Arg288* |
| chr1 | 62602316 | AGATG | A | AF=1e-06 | AQ=58 | frameshift variant | p.Gly291fs |
| chr1 | 62602346 | G | A | AF=1e-06 | AQ=35 | stop gained | p.Trp299* |
| chr1 | 62602373 | GC | G | AF=2e-06 | AQ=44 | frameshift variant | p.Asp310fs |
| chr1 | 62602381 | G | A | AF=3e-06 | AQ=48 | splice donor variant & intron variant | c.931+1G*>*A |
| chr1 | 62604017 | CTAAT | C | AF=7e-06 | AQ=54 | frameshift variant | p.Asn328fs |
| chr1 | 62604031 | C | T | AF=6e-06 | AQ=52 | stop gained | p.Arg332* |
| chr1 | 62604037 | G | T | AF=1e-06 | AQ=48 | stop gained | p.Glu334* |
| chr1 | 62604170 | AT | A | AF=1e-06 | AQ=45 | frameshift variant | p.Leu379fs |
| chr1 | 62604236 | G | T | AF=5e-06 | AQ=47 | splice donor variant & intron variant | c.1198+1G*>*T |
| chr1 | 62604669 | AC | A | AF=1e-06 | AQ=48 | frameshift variant | p.Leu413fs |

**Supplementary Table 6 | *LPA* Predicted Loss-of-Function Variants. Allele frequency is estimated by UK Biobank.**

| **Chromosome** | **Position** | **Reference Allele** | **Alternate Allele** | **Allele Frequency** | **Allele Quality** | **Variant Type** | **Mutation** |
| --- | --- | --- | --- | --- | --- | --- | --- |
| chr6 | 160532530 | C | G | AF=2e-06 | AQ=45 | splice donor variant & intron variant | c.5961+1G*>*C |
| chr6 | 160537854 | C | A | AF=1e-06 | AQ=37 | splice donor variant & intron variant | c.5842+1G*>*T |
| chr6 | 160537907 | G | C | AF=2e-06 | AQ=44 | stop gained | p.Tyr1930* |
| chr6 | 160540053 | T | A | AF=4e-06 | AQ=44 | stop gained | p.Lys1909* |
| chr6 | 160541134 | A | AC | AF=1e-06 | AQ=41 | frameshift variant | p.Val1856fs |
| chr6 | 160541136 | C | T | AF=1.9e-05 | AQ=50 | stop gained | p.Trp1855* |
| chr6 | 160541183 | T | C | AF=3.1e-05 | AQ=45 | splice acceptor variant & intron variant | c.5520-2A*>*G |
| chr6 | 160542686 | A | G | AF=2e-06 | AQ=39 | splice donor variant & intron variant | c.5519+2T*>*C |
| chr6 | 160542687 | C | T | AF=3e-06 | AQ=40 | splice donor variant & intron variant | c.5519+1G*>*A |
| chr6 | 160542714 | C | T | AF=1e-06 | AQ=43 | stop gained | p.Trp1831* |
| chr6 | 160542715 | C | T | AF=2e-06 | AQ=39 | stop gained | p.Trp1831* |
| chr6 | 160542785 | TC | T | AF=1e-06 | AQ=39 | frameshift variant | p.Lys1808fs |
| chr6 | 160547787 | A | G | AF=1e-06 | AQ=39 | splice donor variant & intron variant | c.5304+2T*>*C |
| chr6 | 160547789 | AT | A | AF=6e-06 | AQ=52 | frameshift variant & splice region variant | p.Asn1768fs |
| chr6 | 160547854 | G | A | AF=1e-06 | AQ=52 | stop gained | p.Gln1747* |
| chr6 | 160547903 | GC | G | AF=2e-06 | AQ=45 | frameshift variant | p.Gly1730fs |
| chr6 | 160548544 | CTG | C | AF=3.9e-05 | AQ=46 | frameshift variant | p.Thr1696fs |
| chr6 | 160548552 | G | C | AF=0.000189 | AQ=54 | stop gained | p.Ser1694* |
| chr6 | 160548559 | G | A | AF=5e-06 | AQ=49 | stop gained | p.Arg1692* |
| chr6 | 160548581 | CCTGA | C | AF=2e-06 | AQ=50 | frameshift variant | p.Ile1683fs |
| chr6 | 160548661 | T | C | AF=0.002261 | AQ=58 | splice acceptor variant & intron variant | c.4974-2A*>*G |
| chr6 | 160556024 | C | T | AF=1e-06 | AQ=38 | splice donor variant & intron variant | c.4973+1G*>*A |
| chr6 | 160556070 | A | AG | AF=1e-06 | AQ=40 | frameshift variant | p.Met1643fs |
| chr6 | 160556125 | G | A | AF=5.9e-05 | AQ=51 | stop gained | p.Arg1625* |
| chr6 | 160556129 | A | AC | AF=1e-06 | AQ=47 | frameshift variant | p.Ser1623fs |
| chr6 | 160557521 | C | T | AF=1e-06 | AQ=50 | stop gained | p.Trp1561* |
| chr6 | 160577135 | C | T | AF=0.000212 | AQ=56 | splice donor variant & intron variant | c.4631+1G*>*A |
| chr6 | 160577138 | AT | A | AF=1e-06 | AQ=43 | frameshift variant | p.Asn1543fs |
| chr6 | 160577236 | G | A | AF=6e-06 | AQ=49 | stop gained | p.Arg1511* |
| chr6 | 160577237 | A | T | AF=2.1e-05 | AQ=46 | stop gained | p.Tyr1510* |
| chr6 | 160577258 | G | T | AF=1.1e-05 | AQ=43 | stop gained | p.Tyr1503* |
| chr6 | 160577296 | C | T | AF=1.1e-05 | AQ=47 | splice acceptor variant & intron variant | c.4472-1G*>*A |
| chr6 | 160578589 | C | A | AF=1.9e-05 | AQ=45 | stop gained | p.Glu1469* |
| chr6 | 160578604 | G | A | AF=2e-06 | AQ=48 | stop gained | p.Arg1464* |
| chr6 | 160585045 | C | T | AF=0.037216 | AQ=62 | splice donor variant & intron variant | c.4289+1G*>*A |
| chr6 | 160585139 | GT | G | AF=1.3e-05 | AQ=45 | frameshift variant | p.Thr1399fs |
| chr6 | 160585146 | G | A | AF=2e-05 | AQ=52 | stop gained | p.Arg1397* |
| chr6 | 160585161 | CA | C | AF=1e-06 | AQ=44 | frameshift variant | p.Asp1392fs |
| chr6 | 160585167 | G | A | AF=1.6e-05 | AQ=49 | stop gained | p.Arg1390* |
| chr6 | 160585206 | C | G | AF=2.1e-05 | AQ=54 | splice acceptor variant & intron variant | c.4130-1G*>*C |
| chr6 | 160585206 | C | T | AF=1e-06 | AQ=48 | splice acceptor variant & intron variant | c.4130-1G*>*A |
| chr6 | 160586448 | C | A | AF=7e-06 | AQ=51 | splice donor variant & intron variant | c.4129+1G*>*T |
| chr6 | 160586448 | C | T | AF=1e-06 | AQ=43 | splice donor variant & intron variant | c.4129+1G*>*A |
| chr6 | 160586463 | AG | A | AF=3e-06 | AQ=42 | frameshift variant | p.Leu1372fs |
| chr6 | 160586540 | G | T | AF=3e-06 | AQ=48 | stop gained | p.Cys1346* |
| chr6 | 160586632 | T | C | AF=5e-06 | AQ=39 | splice acceptor variant & intron variant | c.3948-2A*>*G |
| chr6 | 160589551 | A | G | AF=2e-06 | AQ=43 | splice donor variant & intron variant | c.3947+2T*>*C |
| chr6 | 160589564 | G | C | AF=1e-06 | AQ=35 | stop gained | p.Tyr1312* |
| chr6 | 160589585 | C | T | AF=2e-06 | AQ=43 | stop gained | p.Trp1305* |
| chr6 | 160589653 | G | A | AF=6e-06 | AQ=47 | stop gained | p.Arg1283* |
| chr6 | 160590943 | C | T | AF=2.7e-05 | AQ=47 | splice donor variant & intron variant | c.3787+1G*>*A |
| chr6 | 160590971 | C | CA | AF=1e-06 | AQ=41 | frameshift variant | p.Ala1254fs |
| chr6 | 160590982 | G | T | AF=4e-06 | AQ=42 | stop gained | p.Ser1250* |
| chr6 | 160591040 | TG | T | AF=1e-06 | AQ=47 | frameshift variant | p.Met1231fs |
| chr6 | 160593966 | AT | A | AF=5e-06 | AQ=48 | frameshift variant | p.Tyr1207fs |
| chr6 | 160593990 | C | T | AF=1e-06 | AQ=43 | stop gained | p.Trp1199* |
| chr6 | 160593991 | C | T | AF=1e-06 | AQ=40 | stop gained | p.Trp1199* |
| chr6 | 160594011 | C | T | AF=1e-06 | AQ=36 | stop gained | p.Trp1192* |
| chr6 | 160594051 | G | T | AF=3e-06 | AQ=40 | stop gained | p.Ser1179* |
| chr6 | 160594053 | GC | G | AF=1e-06 | AQ=44 | frameshift variant | p.Gly1178fs |
| chr6 | 160594058 | G | A | AF=3e-06 | AQ=48 | stop gained | p.Arg1177* |
| chr6 | 160594070 | C | A | AF=5e-06 | AQ=43 | stop gained | p.Gly1173* |
| chr6 | 160594119 | T | A | AF=7e-06 | AQ=48 | splice acceptor variant & intron variant | c.3470-2A*>*T |
| chr6 | 160595352 | A | G | AF=2e-06 | AQ=46 | splice donor variant & intron variant | c.3469+2T*>*C |
| chr6 | 160595353 | C | A | AF=1e-06 | AQ=46 | splice donor variant & intron variant | c.3469+1G*>*T |
| chr6 | 160595447 | AG | A | AF=1.1e-05 | AQ=46 | frameshift variant | p.Cys1126fs |
| chr6 | 160595454 | C | T | AF=1.1e-05 | AQ=40 | stop gained | p.Trp1123* |
| chr6 | 160595525 | TG | T | AF=4e-06 | AQ=40 | frameshift variant | p.Arg1100fs |
| chr6 | 160595537 | T | C | AF=5e-06 | AQ=45 | splice acceptor variant & intron variant | c.3288-2A*>*G |
| chr6 | 160599498 | A | G | AF=3e-06 | AQ=42 | splice donor variant & intron variant | c.3287+2T*>*C |
| chr6 | 160599499 | C | T | AF=1e-05 | AQ=47 | splice donor variant & intron variant | c.3287+1G*>*A |
| chr6 | 160599499 | C | G | AF=6e-06 | AQ=42 | splice donor variant & intron variant | c.3287+1G*>*C |
| chr6 | 160599508 | G | T | AF=2e-06 | AQ=31 | stop gained | p.Tyr1093* |
| chr6 | 160599561 | G | A | AF=1e-06 | AQ=46 | stop gained | p.Gln1076* |
| chr6 | 160599600 | G | A | AF=1.9e-05 | AQ=52 | stop gained | p.Arg1063* |
| chr6 | 160599609 | G | A | AF=4.3e-05 | AQ=52 | stop gained | p.Gln1060* |
| chr6 | 160599619 | GT | G | AF=1e-06 | AQ=44 | frameshift variant | p.Tyr1056fs |
| chr6 | 160599655 | CA | C | AF=1e-06 | AQ=39 | frameshift variant | p.Leu1044fs |
| chr6 | 160599660 | C | T | AF=1e-06 | AQ=42 | splice acceptor variant & intron variant | c.3128-1G*>*A |
| chr6 | 160600923 | C | CA | AF=4e-06 | AQ=47 | frameshift variant | p.Glu1041fs |
| chr6 | 160600998 | G | A | AF=1.1e-05 | AQ=51 | stop gained | p.Arg1016* |
| chr6 | 160601003 | AG | A | AF=1.9e-05 | AQ=43 | frameshift variant | p.Leu1014fs |
| chr6 | 160601006 | TTG | T | AF=1.9e-05 | AQ=51 | frameshift variant | p.Cys1012fs |
| chr6 | 160601008 | G | T | AF=5e-06 | AQ=40 | stop gained | p.Cys1012* |
| chr6 | 160601076 | G | A | AF=4e-06 | AQ=46 | stop gained | p.Arg990* |
| chr6 | 160601095 | GC | G | AF=2e-06 | AQ=38 | frameshift variant & splice region variant | p.Gly983fs |
| chr6 | 160601099 | C | G | AF=2e-06 | AQ=39 | splice acceptor variant & intron variant | c.2946-1G*>*C |
| chr6 | 160605045 | C | T | AF=4e-05 | AQ=49 | splice donor variant & intron variant | c.2945+1G*>*A |
| chr6 | 160605045 | C | A | AF=3e-06 | AQ=47 | splice donor variant & intron variant | c.2945+1G*>*T |
| chr6 | 160605054 | G | T | AF=2e-06 | AQ=41 | stop gained | p.Tyr979* |
| chr6 | 160605099 | C | T | AF=1.3e-05 | AQ=39 | stop gained | p.Trp964* |
| chr6 | 160605100 | C | T | AF=2e-06 | AQ=36 | stop gained | p.Trp964* |
| chr6 | 160605103 | GC | G | AF=2e-06 | AQ=32 | frameshift variant | p.Ala963fs |
| chr6 | 160605206 | C | T | AF=2.3e-05 | AQ=50 | splice acceptor variant & intron variant | c.2786-1G*>*A |
| chr6 | 160606475 | A | G | AF=5e-06 | AQ=48 | splice donor variant & intron variant | c.2785+2T*>*C |
| chr6 | 160606531 | CG | C | AF=2.1e-05 | AQ=48 | frameshift variant | p.Val911fs |
| chr6 | 160606578 | C | T | AF=3e-06 | AQ=44 | stop gained | p.Trp895* |
| chr6 | 160606635 | C | CTGCAG | AF=2e-06 | AQ=39 | frameshift variant | p.Arg876fs |
| chr6 | 160611561 | C | T | AF=0.000242 | AQ=49 | splice donor variant & intron variant | c.2603+1G*>*A |
| chr6 | 160611578 | CT | C | AF=5e-06 | AQ=33 | frameshift variant | p.Glu863fs |
| chr6 | 160611613 | G | T | AF=1.9e-05 | AQ=45 | stop gained | p.Ser851* |
| chr6 | 160611662 | G | A | AF=5.3e-05 | AQ=48 | stop gained | p.Arg835* |
| chr6 | 160611663 | A | C | AF=1e-06 | AQ=37 | stop gained | p.Tyr834* |
| chr6 | 160611695 | G | A | AF=1e-06 | AQ=43 | stop gained | p.Gln824* |
| chr6 | 160650337 | C | T | AF=3.2e-05 | AQ=50 | splice donor variant & intron variant | c.209+1G*>*A |
| chr6 | 160650389 | G | T | AF=1e-06 | AQ=42 | stop gained | p.Ser53* |
| chr6 | 160650392 | C | T | AF=6e-06 | AQ=48 | stop gained | p.Trp52* |
| chr6 | 160650427 | G | C | AF=1e-06 | AQ=44 | stop gained | p.Tyr40* |
| chr6 | 160650438 | G | A | AF=3.7e-05 | AQ=52 | stop gained | p.Arg37* |
| chr6 | 160650486 | G | A | AF=1e-06 | AQ=46 | stop gained | p.Gln21* |
| chr6 | 160664168 | G | C | AF=1e-06 | AQ=39 | stop gained & splice region variant | p.Ser16* |

**Supplementary Table 7 | *ASGR1* Predicted Loss-of-Function Variants. Allele frequency is estimated by UK Biobank.**

| **Chromosome** | **Position** | **Reference Allele** | **Alternate Allele** | **Allele Frequency** | **Allele Quality** | **Variant Type** | **Mutation** |
| --- | --- | --- | --- | --- | --- | --- | --- |
| chr17 | 7173785 | TC | T | AF=1e-06 | AQ=42 | frameshift variant | p.Gly250fs |
| chr17 | 7173795 | TG | T | AF=2e-06 | AQ=47 | frameshift variant | p.His247fs |
| chr17 | 7173960 | C | G | AF=1e-06 | AQ=35 | splice donor variant & intron variant | c.701+1G*>*C |
| chr17 | 7174256 | A | AC | AF=4e-06 | AQ=50 | frameshift variant | p.Val159fs |
| chr17 | 7174288 | GC | G | AF=1e-06 | AQ=44 | frameshift variant & splice region variant | p.Gly148fs |
| chr17 | 7174461 | C | T | AF=3e-06 | AQ=49 | splice acceptor variant & intron variant | c.356-1G*>*A |
| chr17 | 7174462 | T | C | AF=7.7e-05 | AQ=48 | splice acceptor variant & intron variant | c.356-2A*>*G |
| chr17 | 7176829 | C | G | AF=1e-06 | AQ=41 | splice donor variant & intron variant | c.355+1G*>*C |
| chr17 | 7176830 | C | CT | AF=6e-06 | AQ=45 | frameshift variant & splice region variant | p.Asp119fs |
| chr17 | 7176833 | C | A | AF=6e-06 | AQ=39 | stop gained | p.Glu118* |
| chr17 | 7176847 | TG | T | AF=1e-06 | AQ=35 | frameshift variant | p.Gln113fs |
| chr17 | 7176903 | T | C | AF=7e-06 | AQ=41 | splice acceptor variant & intron variant | c.284-2A*>*G |
| chr17 | 7176980 | C | T | AF=7e-06 | AQ=44 | splice donor variant & intron variant | c.283+1G*>*A |
| chr17 | 7176981 | C | A | AF=1.9e-05 | AQ=42 | stop gained & splice region variant | p.Gly95* |
| chr17 | 7176997 | C | CT | AF=3e-06 | AQ=41 | frameshift variant | p.Ser92fs |
| chr17 | 7177005 | G | A | AF=1e-06 | AQ=37 | stop gained | p.Gln87* |
| chr17 | 7177208 | A | G | AF=1.9e-05 | AQ=48 | splice donor variant & intron variant | c.187+2T*>*C |
| chr17 | 7177300 | G | A | AF=1e-06 | AQ=35 | stop gained | p.Gln33* |
| chr17 | 7178504 | C | CTGAT | AF=5e-06 | AQ=43 | frameshift variant | p.Leu21fs |
| chr17 | 7178539 | G | A | AF=5e-06 | AQ=47 | stop gained | p.Gln9* |
| chr17 | 7178634 | C | G | AF=1e-06 | AQ=39 | splice acceptor variant & intron variant | c.-70-1G*>*C |

**Supplementary Table 8 | A summary of biomarker changes reported in clinical trials for inhibitory drugs targeting *PCSK9*: MK-0616, Alirocumab, Evolocumab, Bococizumab, and Inclisiran**

| ***PCSK9* Trial Summaries** | |
| --- | --- |
| Drug | MK-0616 |
| Drug Type | Oral Macrocyclic Peptide Inhibitor |
| Number of Trials | 1 |
| LDL-C | -40% to -60% |
| ApoB | -32% to -52% |
| Drug | Alirocumab |
| Drug Type | Monoclonal Antibody |
| Number of Trials | 5 |
| LDL-C | -10% to -68% |
| HDL-C | +6% to +12% |
| Total Cholesterol | -6% to -43% |
| Triglycerides | +5% to -17%  *Only one study showed increase in triglycerides |
| ApoA | +1% to +14% |
| ApoB | -11% to -53% |
| Lipoprotein(A) | -7% to -29% |
| Drug | Evolocumab |
| Drug Type | Monoclonal Antibody |
| Number of Trials | 17 |
| LDL-C | -16% to -83% |
| HDL-C | +4% to +12% |
| Total Cholesterol | -28% to -45% |
| Triglycerides | -2% to -15% |
| ApoA |  |
| ApoB | -11% to -52% |
| Lipoprotein(A) | -0.67% to -37% |
| Drug | Bococizumab |
| Drug Type | Monoclonal Antibody |
| Number of Trials | 2 |
| LDL-C | -14% to -66% |
| HDL-C | +3% to +12% |
| Total Cholesterol | -34% to -50% |
| Triglycerides | -8% to -27% |
| Lipoprotein(A) | -46% to -60% |
| Drug | Inclisiran |
| Drug Type | Small Interfering RNA |
| Number of Trials | 5 |
| LDL-C | -20% to -56% |
| HDL-C | -20% to +10%  *Only one trial reported negative HDL-C change |
| Total Cholesterol | -17% to -33% |
| Triglycerides | -20% |
| ApoA | +8% to -14% |
| ApoB | -22% to -40% |
| Lipoprotein(A) | -11% to -25% |
| **Individual *PCSK9* Trials** | |
| Trial | A Study of the Efficacy and Safety of MK-0616 (Oral PCSK9 Inhibitor) in Adults With Hypercholesterolemia (MK-0616-008) |
| Drug | MK-0616 |
| Drug Type | Oral Macrocyclic Peptide Inhibitor |
| Biomarker Changes | -40% to -60% LDL-C, -32% to -52% apoB |
| Identifier | NCT05261126 |
| Trial | A Study of Alirocumab in Participants With Autosomal Dominant Hypercholesterolemia (ADH) and Gain-of-Function Mutations (GOFm) of the Proprotein Convertase Subtilisin Kexin 9 (PCSK9) Gene or Loss-of-Function Mutations (LOFm) of the Apolipoprotein (Apo) B Gene |
| Drug | Alirocumab |
| Drug Type | Monoclonal Antibody |
| Biomarker Changes | -48% to -62% LDL-C, -47% to 53% apoB, -29% to -37% total cholesterol |
| Identifier | NCT01604824 |
| Trial | Study of the Safety and Efficacy of REGN727/SAR236553 in Patients with HeFH Hypercholesterolemia |
| Drug | Alirocumab |
| Drug Type | Monoclonal Antibody |
| Biomarker Changes | -28% to -68% LDL-C, -18% to -43% total cholesterol, +7% to +12 % HDL-C, -4% to -17% triglycerides, -20% to -50% apoB, +1% to +8% apoA, -7% to -23% lpA |
| Identifier | NCT01266876 |
| Trial | Efficacy and Safety of Alirocumab (SAR236553/REGN727)  Versus Placebo on Top of Lipid-Modifying Therapy in Patients with HeFH Not Adequately Controlled with Their Lipid-Modifying Therapy |
| Drug | Alirocumab |
| Drug Type | Monoclonal Antibody |
| Biomarker Changes | -43% to -48% LDL-C, -41% apoB, -31% total cholesterol, -25% lpA, +8% HDL-C, -9% triglycerides, +5% apoA, -21% lpA |
| Identifier | NCT01623115 |
| Trial | An Efficacy and Safety Study of Alirocumab in Children and  Adolescents with HoFH |
| Drug | Alirocumab |
| Drug Type | Monoclonal Antibody |
| Biomarker Changes | -10% LDL-C, -11% apoB, -6% total cholesterol, -5% lpA, +8% HDL-C, +14% apoA, +5% triglycerides |
| Identifier | NCT03510715 |
| Trial | Reduction of LDL-C with PCSK9 Inhibition in HeFH Disorder Study |
| Drug | Evolocumab |
| Drug Type | Monoclonal Antibody |
| Biomarker Changes | -42% to -55% LDL-C, -30% to -40% apoB, -33% to -42% total cholesterol |
| Identifier | NCT01375751 |
| Trial | LAPLACE-TIMI 57: LDL-C Assessment with PCSK9 monoclonal Antibody Inhibition Combined with Statin therapy |
| Drug | Evolocumab |
| Drug Type | Monoclonal Antibody |
| Biomarker Changes | -39% to -63% LDL-C, -28% to -50% apoB, -28% to -45% total cholesterol, |
| Identifier | NCT01380730 |
| Trial | Trial Assessing Long-Term Use of PCSK9 Inhibition in Subjects with Genetic LDL Disorders |
| Drug | Evolocumab |
| Drug Type | Monoclonal Antibody |
| Biomarker Changes | -32% to -50% LDL-C, -0.67% to -29% lpA, -15% to -48% apoB, |
| Identifier | NCT01624142 |
| Trial | Monoclonal Antibody Against PCSK9 to Reduce Elevated LDL-C in Adults Currently Not Receiving Drug Therapy for Easing Lipid Levels |
| Drug | Evolocumab |
| Drug Type | Monoclonal Antibody |
| Biomarker Changes | -39% to -50% LDL-C, -32% to -45% apoB |
| Identifier | NCT01375777 |
| Trial | Goal Achievement After Utilizing an Anti-PCSK9 Antibody in Statin Intolerant Subjects |
| Drug | Evolocumab |
| Drug Type | Monoclonal Antibody |
| Biomarker Changes | -40% to -50% LDL-C, -33% to -42% apoB |
| Identifier | NCT01375764 |
| Trial | Goal Achievement After Utilizing an Anti-PCSK9 Antibody in Statin Intolerant Subjects -2 |
| Drug | Evolocumab |
| Drug Type | Monoclonal Antibody |
| Biomarker Changes | -52% to -56% LDL-C, -43% to -45% ApoB -23% to -26% Lp(A), -2% to -3% TG, +5% to +6% HDL-C |
| Identifier | NCT01763905 |
| Trial | Goal Achievement After Utilizing an Anti-PCSK9 Antibody in Statin Intolerant Subjects-3 |
| Drug | Evolocumab |
| Drug Type | Monoclonal Antibody |
| Biomarker Changes | -54% LDL-C, -38% total cholesterol, -45% apoB, -22% lpA, -5% triglycerides, +7% HDL-C |
| Identifier | NCT01984424 |
| Trial | Goal Achievement After Utilizing an Anti-PCSK9 Antibody in Statin Intolerant Subjects-4 |
| Drug | Evolocumab |
| Drug Type | Monoclonal Antibody |
| Biomarker Changes | -59% LDL-C, -39% total cholesterol, -48% apoB, -37% lpA, -2% triglycerides, +11% HDL-C |
| Identifier | NCT02634580 |
| Trial | Trial Evaluating PCSK9 Antibody in Subjects with LDL Receptor Abnormalities |
| Drug | Evolocumab |
| Drug Type | Monoclonal Antibody |
| Biomarker Changes | -16% to -23% LDL-C, -15% apoB, -9% Lp(A) |
| Identifier | NCT01588496 |
| Trial | Effects of PCSK9 Inhibition on Arterial Wall Inflammation in Patients with Elevated Lipoprotein(a) (Lp(a)) |
| Drug | Evolocumab |
| Drug Type | Monoclonal Antibody |
| Biomarker Changes | -59% LDL-C, -12% lpA, -48% apoB |
| Identifier | NCT02729025 |
| Trial | Trial Assessing Efficacy, Safety, and Tolerability of Proprotein  Convertase Subtilisin/Kexin Type 9 (PCSK9) Inhibition in  Pediatric Subjects with Genetic LDL Disorders |
| Drug | Evolocumab |
| Drug Type | Monoclonal Antibody |
| Biomarker Changes | -44% to -77% LDL-C, -35% apoB, |
| Identifier | NCT02392559 |
| Trial | Durable Effect of PCSK9 Antibody Compared with placebo Study |
| Drug | Evolocumab |
| Drug Type | Monoclonal Antibody |
| Biomarker Changes | -50% LDL-C, -32% total cholesterol, -41% apoB, -27% lpA, -2% triglycerides, +5% HDL-C |
| Identifier | NCT01516879 |
| Trial | Effects on Lipoprotein Metabolism from PCSK9 Inhibition Utilizing a Monoclonal Antibody |
| Drug | Evolocumab |
| Drug Type | Monoclonal Antibody |
| Biomarker Changes | -57% to -83% LDL-C |
| Identifier | NCT02189837 |
| Trial | Reduction of LDL-C with PCSK9 Inhibition in HeFH Disorder Study-2 |
| Drug | Evolocumab |
| Drug Type | Monoclonal Antibody |
| Biomarker Changes | -55% to -61% LDL-C, -49% to -52% apoB, -25% lpA, -9% to -13% triglycerides, +6% to +8% HDL-C |
| Identifier | NCT01763918 |
| Trial | Monoclonal Antibody Against PCSK9 to Reduce Elevated  LDL-C in Subjects Currently Not Receiving Drug Therapy for Easing Lipid Levels-2 |
| Drug | Evolocumab |
| Drug Type | Monoclonal Antibody |
| Biomarker Changes | -56% LDL-C, -49% apoB, -17 to -20% lpA, -8% to -15% triglycerides, +4% HDL-C |
| Identifier | NCT01763827 |
| Trial | Evolocumab Compared to LDL-C Apheresis in Patients Receiving LDL-C Apheresis Prior to Study Enrollment |
| Drug | Evolocumab |
| Drug Type | Monoclonal Antibody |
| Biomarker Changes | -50% LDL-C, -35% total cholesterol |
| Identifier | NCT02585895 |
| Trial | Safety, Tolerability, and Efficacy on Low-Density Lipoprotein Cholesterol (LDL-C) of Evolocumab in Participants with Human Immunodeficiency Virus (HIV) and Hyperlipidemia/Mixed Dyslipidemia |
| Drug | Evolocumab |
| Drug Type | Monoclonal Antibody |
| Biomarker Changes | -55% LDL-C, -45% apoB, -35% total cholesterol, -16% lpA, -9% triglycerides, +11% HDL-C |
| Identifier | NCT02833844 |
| Trial | Dose Ranging Study Of Bococizumab (PF-04950615; RN316) In Hypercholesterolemic Japanese Subjects |
| Drug | Bococizumab |
| Drug Type | Monoclonal Antibody |
| Biomarker Changes | -49% to -66% LDL-C, -34% to -50% total cholesterol, +5% to +12% HDL-C, -13% to -27% triglycerides, -46% to -60% lpA |
| Identifier | NCT02055976 |
| Trial | A Multiple Dose Study Of PF-04950615 (RN316) In Subjects On Maximum Doses Of Statins |
| Drug | Bococizumab |
| Drug Type | Monoclonal Antibody |
| Biomarker Changes | -14% to -44% LDL-C, +3% to +5% HDL-C, -9% to -32%, -8% to -11% triglycerides |
| Identifier | NCT01350141 |
| Trial | Inclisiran for Subjects With ASCVD or ASCVD-Risk Equivalents and Elevated Low-density Lipoprotein Cholesterol (ORION-11) |
| Drug | Inclisiran Sodium |
| Drug Type | Small Interfering RNA |
| Biomarker Changes | -49% LDL-C, -28% TC, -38% apoB |
| Identifier | NCT03400800 |
| Trial | Inclisiran for Participants with Atherosclerotic Cardiovascular  Disease and Elevated Low-density Lipoprotein Cholesterol |
| Drug | Inclisiran Sodium |
| Drug Type | Small Interfering RNA |
| Biomarker Changes | -56% LDL-C, -33% TC, -44% ApoB, |
| Identifier | NCT03399370 |
| Trial | Trial to Evaluate the Effect of Inclisiran Treatment on Low-Density Lipoprotein Cholesterol (LDL-C) in Subjects with HeFH |
| Drug | Inclisiran |
| Drug Type | Small Interfering RNA |
| Biomarker Changes | -41% LDL-C, -25% TC, -33% ApoB, |
| Identifier | NCT03397121 |
| Trial | A Study of ALN-PCSSC in Participants With Homozygous Familial Hypercholesterolemia (HoFH) (ORION-2) |
| Drug | Inclisiran |
| Drug Type | Small Interfering RNA |
| Biomarker Changes | -20% LDL-C, -18% TC, -20% TG, -20% HDL-C, -14% apoA, -25% apoB, -11% lpA |
| Note: | Only looked at 4 participants |
| Identifier | NCT02963311 |
| Trial | Trial to Evaluate the Effect of ALN-PCSSC Treatment on Low Density Lipoprotein Cholesterol (LDL-C) |
| Drug | Inclisiran |
| Drug Type | Small Interfering RNA |
| Biomarker Changes | -27% to -52% LDL-C, -17% to -33% TC, +4% to +10% HDL-C, -22% to -40% ApoB, +2% to +8% ApoA, -14% to -25% Lp(A) |
| Identifier | NCT02597127 |
| Trial | Open-Label Extension of Study R727-CL-1003 (NCT01266876) to Evaluate the Long-Term Safety and Efficacy of Alirocumab (REGN727) in Participants with HeFH |
| Drug | Alirocumab |
| Drug Type | Monoclonal Antibody |
| Biomarker Changes | -65% LDL-C, -50% ApoB, -41% TC, -29% Lp(A), +6% to +8% HDL-C, -0.9% to -6% TG, +4% to +11% ApoA |
| Identifier | NCT01576484 |

**Supplementary Table 9 | A summary of biomarker changes reported in clinical trials for inhibitory drugs targeting *APOC3*: Volanesorsen, Olezarsen (AKCEA-APOCIII-LRx), and ARO-APOC3**

| ***APOC3* Trial Summaries** | | |
| --- | --- | --- |
| Drug | | Volanesorsen |
| Drug Type | | Antisense Oligonucleotide |
| Number of Trials | | 5 |
| LDL-C | | 0% to -21% |
| HDL-C | | +26% to +61% |
| Triglycerides | | -31% to -76% |
| ApoB | | -20% |
| Drug | | Olezarsen (AKCEA-APOCIII-LRx) |
| Drug Type | | N-acetyl galactosamine-conjugated (GalNAc3) ASO |
| Number of Trials | | 2 |
| LDL-C | | -21% to +27%  *Researchers noted cause for LDL-C increase |
| HDL-C | | +11% to +75% |
| Total Cholesterol | | -2% to -16% |
| Triglycerides | | -23% to -70% |
| ApoA | | +5% top +18% |
| ApoB | | 0% to -30% |
| Drug | | ARO-APOC3 |
| Drug Type | | Small interfering RNA |
| Number of Trials | | 3 |
| LDL-C | | +2% to -25% |
| HDL-C | | +28% to +136% |
| Triglycerides | | -41% to -92% |
| **Individual *APOC3* Trials** | | |
| Trial | The APPROACH Study: A Study of Volanesorsen (Formerly IONIS-APOCIIIRx) in Patients With Familial Chylomicronemia Syndrome | |
| Drug | Volanesorsen | |
| Drug Type | Antisense Oligonucleotide | |
| Biomarker Changes | -76% TG | |
| Identifier | NCT02658175 | |
| Trial | Study of ISIS 678354 (AKCEA-APOCIII-LRx) in Participants With Hypertriglyceridemia and Established Cardiovascular Disease (CVD) | |
| Drug | Olezarsen (AKCEA-APOCIII-LRx) | |
| Drug Type | N-acetyl galactosamine-conjugated (GalNAc3) ASO | |
| Biomarker Changes | -23% to -60% TG, -2% to -12% TC, +2% to +27% LDL-C, +11% to +40% HDL-C, 0% to -17% ApoB, +5% to +18% ApoA | |
| Note: | “The increase in LDL-C in the 10 mg weekly group appeared to be related to changes or discontinuation of in background LDL-C-lowering therapies in some patients.” | |
| Identifier | NCT03385239 | |
| Trial | Safety, Tolerability, PK, and Pharmacodynamics(PD) of IONIS APOCIII-LRx in Healthy Volunteers With Elevated Triglycerides | |
| Drug | Olezarsen (AKCEA-APOCIII-LRx) | |
| Drug Type | N-acetyl galactosamine-conjugated (GalNAc3) ASO | |
| Biomarker Changes | -60% to -70% TG, -5% to -16% TC, -2% to -21% LDL-C, -15% to -30% ApoB, +49% to +75% HDL-C | |
| Identifier | NCT02900027 | |
| Trial | The COMPASS Study: A Study of Volanesorsen (Formally ISIS-APOCIIIRx) in Patients With Hypertriglyceridemia | |
| Drug | Volanesorsen | |
| Drug Type | Antisense Oligonucleotide | |
| Biomarker Changes | -71% TG, +61% HDL-C | |
| Identifier | NCT02300233 | |
| Trial | Safety, Tolerability, and Pharmacokinetic Study of ISIS ApoC-III Rx in Hypertriglyceridemia | |
| Drug | Volanesorsen | |
| Drug Type | Antisense Oligonucleotide | |
| Biomarker Changes | -31% to -70% TG, +26% to +45% HDL-C | |
| Identifier | NCT01529424 | |
| Paper | Antisense-Mediated Lowering of Plasma Apolipoprotein C-III by Volanesorsen Improves Dyslipidemia and Insulin Sensitivity in Type 2 Diabetes | |
| Drug | Volanesorsen | |
| Drug Type | Antisense Oligonucleotide | |
| Biomarker Changes | -69% TG, +42% HDL-C, LDL-C 0%, -20% ApoB | |
| Link | https://doi.org/10.2337/dc16-0126 | |
| Trial | The Approach Open Label Study: A Study of Volanesorsen (Formerly IONIS-APOCIIIRx) in Participants With Familial Chylomicronemia Syndrome | |
| Drug | Volanesorsen | |
| Drug Type | Antisense Oligonucleotide | |
| Biomarker Changes | -34% to -65% TG, | |
| Identifier | NCT02658175 | |
| Trial | Study of ARO-APOC3 in Adults With Mixed Dyslipidemia (MUIR) | |
| Drug | ARO-APOC3 | |
| Drug Type | Small interfering RNA | |
| Biomarker Changes | -49% to -62% TG, +2% to -13% LDL-C, +28% to +45% HDL-C, -37% to -48% Remnant Cholesterol, -6% to -20% ApoB | |
| Identifier | NCT04998201 | |
| Trial | Study of ARO-APOC3 in Healthy Volunteers, Hypertriglyceridemic Patients and Patients With Familial Chylomicronemia Syndrome (FCS) | |
| Drug | ARO-APOC3 | |
| Drug Type | Small interfering RNA | |
| Biomarker Changes | -78% to -92% TG, +71% to +136% HDL-C | |
| Trial | NCT03783377 | |
| Paper | RNA Interference Targeting Apolipoprotein C-III Results in Deep and Prolonged Reductions in Plasma Triglycerides | |
| Drug | ARO-APOC3 | |
| Drug Type | Small interfering RNA | |
| Biomarker Changes | -41% to -64% TG, -12% to -25% LDL-C, +28% to +69% HDL-C, | |
| Link | https://professional.heart.org/-/media/PHD-Files/Meetings/Scientific-Sessions/2019/Sci-News-2019/RNA_Interference_Targeting_Apolipoprotein_C_III_Results_in_Deep_ucm_505213.pdf | |

**Supplementary Table 10 | A summary of biomarker changes reported in clinical trials for inhibitory drugs targeting *ANGPTL3*: Evinacumab,** **Vupanorsen, and ARO-ANG3**

| ***ANGPTL3* Trial Summaries** | |
| --- | --- |
| Drug | Evinacumab |
| Drug Type | Monoclonal Antibody |
| Number of Trials | 5 |
| LDL-C | -9% to -52% |
| HDL-C | +8% to -37% |
| Total Cholesterol | -7% to -60% |
| Triglycerides | -19% to -52%  *Note: only one trial reported positive increase |
| ApoA | +4% to -38% |
| ApoB | -2% to -45% |
| Drug | Vupanorsen |
| Drug Type | N-acetyl galactosamine (GalNAc3)-modified ASO |
| Number of Trials | 4 |
| LDL-C | +5% to -17%  *Note: only one trial reported positive increase |
| HDL-C | 0% to -15% |
| Total Cholesterol | -21% to -41% |
| Triglycerides | -32% to -59% |
| ApoA | -11% to -30% |
| ApoB | -5% to -14% |
| Drug | ARO-ANG3 |
| Drug Type | Small Interfering RNA |
| Number of Trials | 3 |
| LDL-C | -23% to -44% |
| HDL-C | -14% to -37% |
| Triglycerides | -25% to -79% |
| ApoB | -28% to -39% |
| **Individual *ANGPTL3* Trials** | |
| Trial | Study of Evinacumab (REGN1500) in Participants With Persistent Hypercholesterolemia |
| Drug | Evinacumab |
| Drug Type | Monoclonal Antibody |
| Biomarker Changes | -23% to -49% LDL-C, -19% to -43% ApoB, -22% to -46% TC, -32% to -52% TG, -8% to -15% Lp(A) |
| Identifier | NCT03175367 |
| Trial | Evaluate the Efficacy and Safety of Evinacumab in Pediatric Patients With Homozygous Familial Hypercholesterolemia |
| Drug | Evinacumab |
| Drug Type | Monoclonal Antibody |
| Biomarker Changes | -48% LDL-C, -41% ApoB, -49% TC, -37% Lp(A) |
| Identifier | NCT04233918 |
| Trial | Efficacy and Safety of Evinacumab in Patients With Homozygous Familial Hypercholesterolemia |
| Drug | Evinacumab |
| Drug Type | Monoclonal Antibody |
| Biomarker Changes | -47% LDL-C, -41% ApoB, -47% TC, -55% TG, -5% Lp(A) |
| Identifier | NCT03399786 |
| Trial | Safety and Efficacy Following Repeat-Dose of Evinacumab (Anti-ANGPTL3) in Patients With Severe Hypertriglyceridemia (sHTG) at Risk for Acute Pancreatitis |
| Drug | Evinacumab |
| Drug Type | Monoclonal Antibody |
| Biomarker Changes | -35% to -39% TG |
| Identifier | NCT03452228 |
| Trial | Study of REGN1500 in Participants With Homozygous Familial Hypercholesterolemia (HoFH) |
| Drug | Evinacumab |
| Drug Type | Monoclonal Antibody |
| Biomarker Changes | -9% to -52% LDL-C, -2% to -45% ApoB, -7% to -60% TC, +40% to -23% Lp(A), -18% HDL-C, -21% TG, -19% ApoA  *TG, HDL-C, and ApoA are Averages |
| Identifier | NCT02265952 |
| Trial | Study of ISIS 703802 in Participants With Hypertriglyceridemia, Type 2 Diabetes Mellitus, and Nonalcoholic Fatty Liver Disease |
| Drug | Vupanorsen |
| Drug Type | N-acetyl galactosamine (GalNAc3)-modified ASO |
| Biomarker Changes | -36% to -53% TG, -21% to -41% TC, +5% to -11% LDL-C, -0.6% to -6% HDL-C, -6% to -13% ApoB, -11% to -30% ApoA |
| Identifier | NCT03371355 |
| Trial | A Dose-Ranging Study With Vupanorsen (TRANSLATE-TIMI 70) |
| Drug | Vupanorsen |
| Drug Type | N-acetyl galactosamine (GalNAc3)-modified ASO |
| Biomarker Changes | -39% to -58% TG, -5% to -14% ApoB, -2% to -17% LDL-C, |
| Identifier | NCT04516291 |
| Trial | Phase 2 Study of AKCEA-ANGPTL3-LRx (ISIS 703802) in Participants With Familial Chylomicronemia Syndrome (FCS) |
| Drug | Vupanorsen |
| Drug Type | N-acetyl galactosamine (GalNAc3)-modified ASO |
| Biomarker Changes | -32% TG, -30% TC, -10% ApoB, -15% HDL-C, -17% ApoA, -6% LDL-C |
| Identifier | NCT03360747 |
| Trial | Study of AKCEA-ANGPTL3-LRx (ISIS 703802) in Participants With Familial Partial Lipodystrophy (FPL) |
| Drug | Vupanorsen |
| Drug Type | N-acetyl galactosamine (GalNAc3)-modified ASO |
| Biomarker Changes | -59% TG |
| Identifier | NCT03514420 |
| Trial | Study of ARO-ANG3 in Healthy Volunteers and in Dyslipidemic Patients |
| Drug | ARO-ANG3 |
| Drug Type | Small Interfering RNA |
| Biomarker Changes | -61% to -66% TG, -34% to -44% LDL-C, -28% to -39% ApoB, -14% to -37% HDL-C, -35% to -43% TC, -23% to -38% ApoA |
| Link | <https://www.nature.com/articles/s41591-023-02494-2>  Table S4 |
| Abstract | Abstract 15751: Pharmacodynamic Effect of ARO-ANG3, an Investigational RNA Interference Targeting Hepatic Angiopoietin-like Protein 3, in Patients With Hypercholesterolemia |
| Drug | ARO-ANG3 |
| Drug Type | Small Interfering RNA |
| Biomarker Changes | -25% to -43% TG, -23% to -37% LDL-C |
| Link | https://www.ahajournals.org/doi/10.1161/circ.142.suppl_3.15751 |
| Abstract | Reduced Expression of Angiopoietin-Like Protein 3 via RNA Interference with AROANG3 Produces Prolonged Reductions in LDL-C and Triglycerides in Dyslipidemic Patients |
| Drug | ARO-ANG3 |
| Drug Type | Small Interfering RNA |
| Biomarker Changes | -39% to -42% LDL-C, -79% TG |
| Link | https://doi.org/10.1016/j.jacl.2020.05.085 |

**Supplementary Table 11 | A summary of biomarker changes reported in clinical trials for inhibitory drugs targeting *LPA*: Lepodisiran,** **ISIS-APO(a)Rx, and Olpasiran (AMG 890)**

| ***LPA* Trial Summaries** | |
| --- | --- |
| Drug | Lepodisiran |
| Drug Type | Small Interfering RNA |
| Number of Trials | 1 |
| Lipoprotein (A) | -41% to -97% |
| Drug | ISIS-APO(a)Rx |
| Drug Type | Antisense Oligonucleotides |
| Number of Trials | 2 |
| Lipoprotein (A) | -39% to -77% |
| Drug | Olpasiran (AMG 890) |
| Drug Type | Small Interfering RNA |
| Number of Trials | 2 |
| LDL-C | -11% to -17% |
| Lipoprotein (A) | -64% to -97% |
| ApoB | -4% to -9% |
| **Individual *LPA* Trials** | |
| Trial | A Study of LY3819469 in Healthy Participants |
| Drug | Lepodisiran |
| Drug Type | Small Interfering RNA |
| Biomarker Changes | -41% to -97% Lp(A) |
| Link | doi:10.1001/jama.2023.21835 |
| Identifier | NCT04914546 |
| Trial | Safety, Tolerability, Pharmacokinetics, and Pharmacodynamics of ISIS-APO(a)Rx in Participants With High Lipoprotein(a) |
| Drug | ISIS-APO(a)Rx |
| Drug Type | Antisense Oligonucleotides |
| Biomarker Changes | -44% to -71% Lp(A) |
| Identifier | NCT02160899 |
| Paper | Antisense therapy targeting apolipoprotein(a): a randomised, double-blind, placebo-controlled phase 1 study |
| Drug | ISIS-APO(a)Rx |
| Drug Type | Antisense Oligonucleotides |
| Biomarker Changes | -39% to -77% Lp(A) |
| DOI | https://doi.org/10.1016/s0140-6736(15)61252-1 |
| Trial | Olpasiran Trials of Cardiovascular Events And Lipoprotein(a) Reduction - DOSE Finding Study |
| Drug | Olpasiran (AMG 890) |
| Drug Type | Small Interfering RNA |
| Biomarker Changes | -64% to 97% Lp(A), -11% to -17% LDL-C, -4% to -9% ApoB |
| Identifier | NCT04270760 |
| Trial | Safety, Tolerability, Pharmacokinetics and Pharmacodynamics Study of AMG 890 in Subjects With Elevated Plasma Lipoprotein(a) |
| Drug | Olpasiran (AMG 890) |
| Drug Type | Small Interfering RNA |
| Biomarker Changes | -71% to -97% Lp(A) |
| Identifier | NCT03626662 |

**Supplementary Table 12 | A summary of biomarker changes reported in clinical trials for inhibitory drugs targeting *ASGR1*: AMG 529**

| *ASGR1* Trial Summaries | |
| --- | --- |
| Drug | AMG 529 |
| Drug Type | Monoclonal Antibody |
| Number of Trials | 2 |
| LDL-C | +1% to -5% |
| Total Cholesterol | +2% to -3% |
| Alkaline Phosphatase | +251% |
| Individual *ASGR1* Trials | |
| Trial | AMG 529 First in Human Study |
| Drug | AMG 529 |
| Drug Type | Monoclonal Antibody |
| Biomarker Changes | +1 to -5% LDL-C, +2% to -3% TC, +251% Alkaline Phosphatase |
| Identifier | NCT03170193 |
